# COVID-19 in India: State-wise Analysis and Prediction

**DOI:** 10.1101/2020.04.24.20077792

**Authors:** Palash Ghosh, Rik Ghosh, Bibhas Chakraborty

**Affiliations:** Department of Mathematics, Indian Institute of Technology Guwahati, India; Centre for Quantitative Medicine, Duke-NUS Medical School, Singapore; Department of Statistics and Applied Probability, National University of Singapore; Department of Biostatistics and Bioinformatics, Duke University, USA

## Abstract

Coronavirus disease 2019 (COVID-19), a highly infectious disease, was first detected in Wuhan, China, in December 2019. The disease has spread to 212 countries and territories around the world and infected (confirmed) more than three million people. In India, the disease was first detected on 30 January 2020 in Kerala in a student who returned from Wuhan. The total (cumulative) number of confirmed infected people is more than 37000 till now across India (3 May 2020). Most of the research and newspaper articles focus on the number of infected people in the entire country. However, given the size and diversity of India, it may be a good idea to look at the spread of the disease in each state separately, along with the entire country. For example, currently, Maharashtra has more than 10000 confirmed cumulative infected cases, whereas West Bengal has less than 800 confirmed infected cases (1 May 2020). The approaches to address the pandemic in the two states must be different due to limited resources. In this article, we will focus the infected people in each state (restricting to only those states with enough data for prediction) and build three growth models to predict infected people for that state in the next 30 days. The impact of preventive measures on daily infected-rate is discussed for each state.

**Highlights of the Analysis:**

- Data considered for analysis: up to 1 May 2020.
- One model can mislead us. Here, we consider the exponential, the logistic and the SIS models along with daily infection-rate (DIR). We interpret the results jointly from all models rather than individually.
- We expect DIR to be zero or negative to conclude that COVID-19 is not spreading in a state. Even a small positive DIR (say 0.01) indicates virus is spreading in the community. The virus can potentially increase the DIR anytime.
- **<u>Severe</u>:** The states without a decreasing trend in DIR and near exponential growth in active infected cases are Maharashtra, Delhi, Gujarat, Madhya Pradesh, Andhra Pradesh, Uttar Pradesh, and West Bengal.
- **<u>Moderate</u>:** The states with an almost decreasing trend in DIR and non-increasing growth in active infected cases are Tamil Nadu, Rajasthan, Punjab and Bihar.
- **<u>Controlled</u>:** The states with a decreasing trend in DIR and decreasing growth in active infected cases in the last few days are Kerala, Haryana, Jammu and Kashmir, Karnataka, and Telangana.
- States with non-decreasing DIR need to do much more in terms of the preventive measures immediately to combat the COVID-19 pandemic. On the other hand, the states with decreasing DIR can maintain the same status to see the DIR become zero or negative for consecutive 14 days to be able to declare the end of the pandemic.

## Introduction

The world is now facing an unprecedented crisis due to the novel coronavirus, first detected in Wuhan, China, in December 2019 [^1^]. World Health Organization (WHO) defined coronavirus as a family of viruses that range from the common cold to the Middle East Respiratory Syndrome (MERS) coronavirus and the Severe acute respiratory syndrome (SARS) coronavirus [^2^]. Coronaviruses circulate in some wild animals and have the capability to transmit from animals to humans. These viruses can cause respiratory symptoms in humans, along with other symptoms of common cold and fever [^3^]. There are no specific treatments for coronaviruses to date. However, one can avoid infection by maintaining basic personal hygiene and social distancing from infected persons.

WHO declared Coronavirus disease 2019 (COVID-19) as a global pandemic on 11 March 2020 [^4^]. The disease has spread across 212 countries and territories around the world, with a total of more than three million confirmed cases [^5,6^]. In India, the disease was first detected on 30 January 2020 in Kerala in a student who returned from Wuhan [^7,8^]. The total (cumulative) number of confirmed infected people is more than 37000 till date (3 May 2020) across India. The bar chart in Figure 1 shows the daily growth of the COVID-19 cases in India. After the first three cases during 30 January-3 February 2020, there were no more confirmed COVID-19 cases for about a month. The COVID-19 cases appeared again from 2 March 2020 onwards. These cases are related to people who have been evacuated or have arrived from COVID-19 affected countries. From 20 March 2020 onwards, there is an exponential growth in the daily number of COVID-19 cases at pan India level.

**Figure 1:**
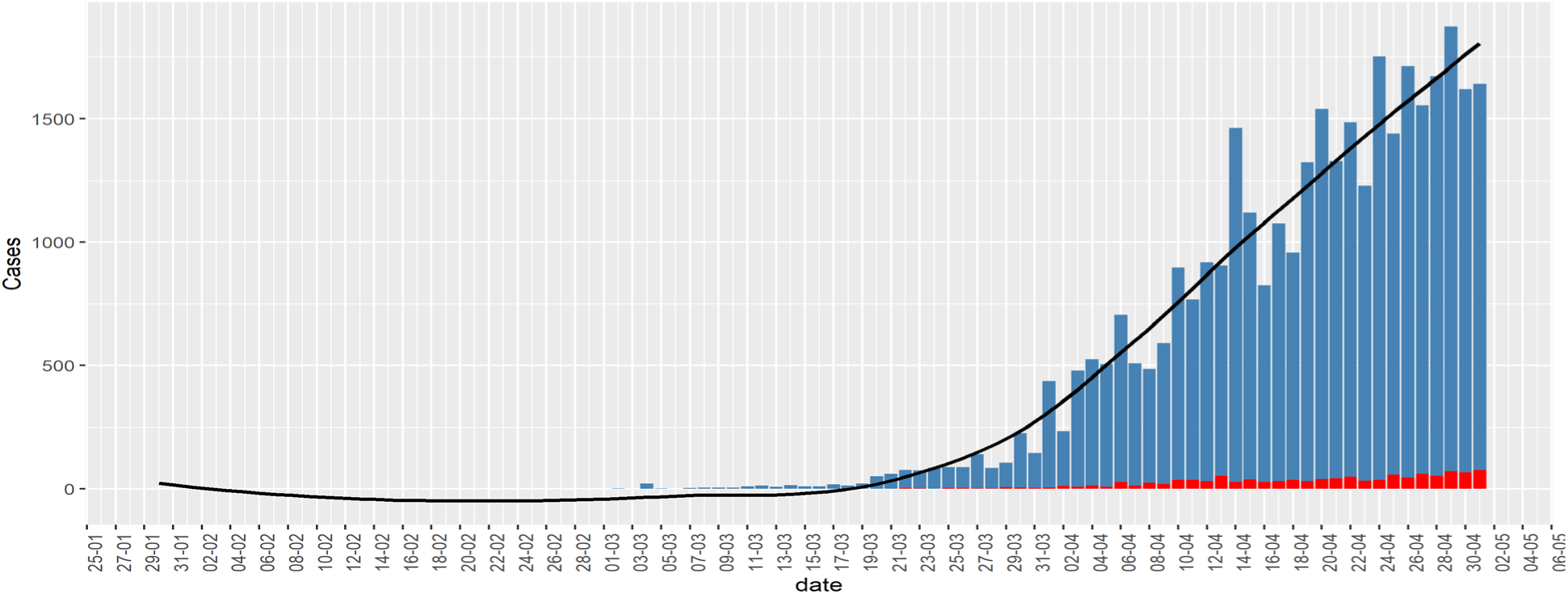
Bar chart of daily infected cases (blue) in India. Red bar denotes death. The black curve is a fitted smooth curve on the daily cases.

There are four stages of COVID-19, depending on the types of virus transmission [^9,10^], as follows. Stage 1: During this stage, a country/region experiences imported infected cases with travel history from virus-hit countries. Stage 2: During this stage, a country/region gets new infections from persons who did not have travel history but came in contact with persons defined in stage 1. Stage 3: This is community transmission; in this period, new infection occurs in a person who has not been in contact with an infected person or anyone with a travel history of virus-hit countries. Stage 4: At this stage, the virus spread is practically uncontrollable, and the country can have many major clusters of infection.

Many news agencies are repeatedly saying or questioning whether India is now at stage 3 [^9,11,12^]. In reality, different Indian states are or will be at various stages of infection at different points in time. Labeling a COVID-19 stage at pan India level is problematic. It will spread misinformation to common people. Those states, which are at stage 3, require more rapid action compared to others. On the other hand, states that are in stages 1 and 2, need to focus on stopping the community-spreading of COVID-19.

In this article, first, we discuss the importance of state-wise consideration, considering all the states together. Then, we will focus on the infected people in each state (considering only those states with enough data for prediction) and build growth models to predict infected people for that state in the next 30 days.

### Why State-wise consideration?

India is a vast country with a geographic area of 3,287,240 square kilometers, and a total population of about 1.3 billion [^13^]. Most of the Indian states are quite large in the geographic area and population. Analyzing coronavirus infection data, considering entire India to be on the same page, may not provide us the right picture. This is so because the first infection, new infection-rate, progression over time, and preventive measures taken by state governments and the common public for each state are different. We need to address each state separately. It will enable the government to utilize the limited available resources optimally. For example, currently, Maharashtra already has more than 10000 confirmed infected cases, whereas West Bengal has less than 800 confirmed cases (1 May 2020). The approaches to addressing the two states must be different due to limited resources. One way to separate the state-wise trajectories is to look at when each state was first Infected.

In Figure 2, we present the first infection date along with the infected person’s travel history in each of the Indian states. All the states and the union territories, except Assam, Tripura, Nagaland, Meghalaya, and Arunachal Pradesh, observed their first confirmed infected case from a person who has travel history from one or more already COVID-19 infected countries. The Indian government imposed a complete ban on international flights to India on 22 March 2020 [^14^]. Figure 1 justifies government action to international flight suspension. Had it been taken earlier, we could have restricted the disease only in few states compared to the current scenario.

**Figure 2:**
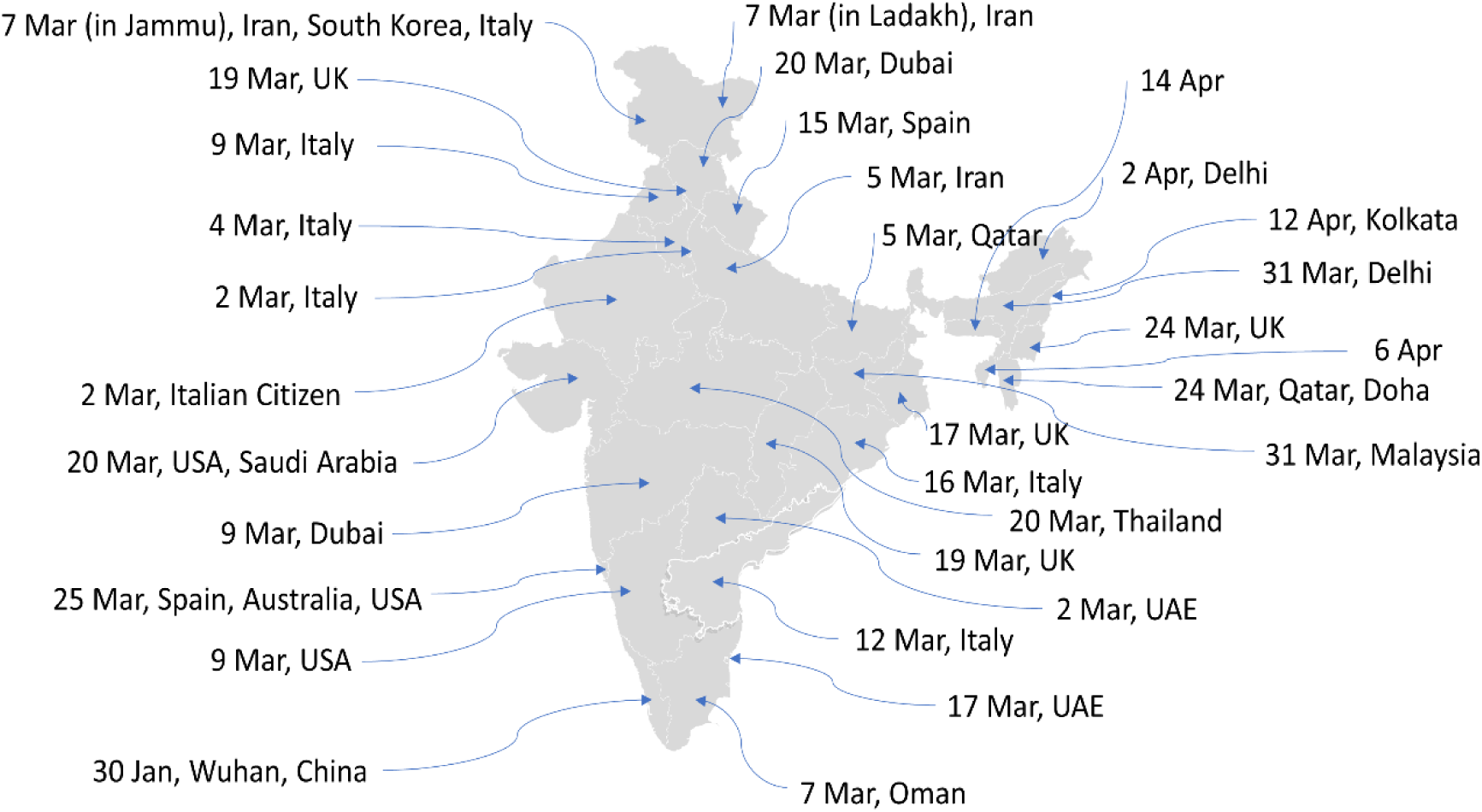
How the first case in each state happened with their travel histories.

Figure 3 shows the line-graph of the cumulative number of infected people in those Indian states having at least 10 total infected people. Currently, Maharashtra, Delhi, Gujarat, Tamil Nadu, Madhya Pradesh, Rajasthan, and Uttar Pradesh are the states where the cumulative number of infected people have crossed the 2000 mark, with Maharashtra having more than 10000 cases. Kerala, the first state to have the COVID-19 confirmed case, seems to have restricted the growth-rate under control. There are few states with cumulative infected people number in the range of 500-1500. Depending on how those states strictly follow the preventive measures, we may see a rise in the confirmed cases.

**Figure 3.**
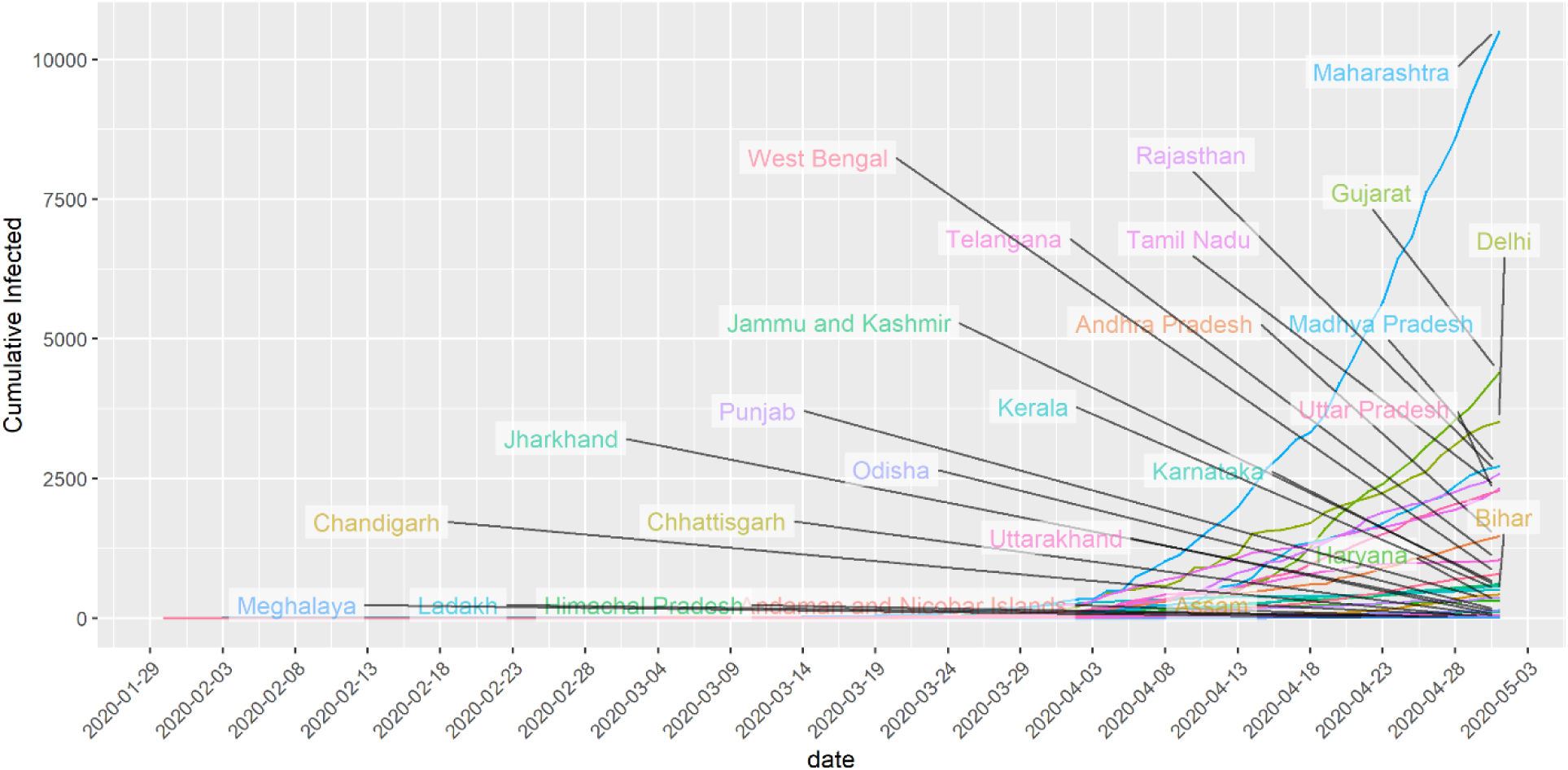
Showing the cumulative infected people in states with at least 10 infected cases.

## Preventive Measures

Below we list the major preventive measures taken by the Indian Government [^15^].

**Table 1:** List of major preventive measures taken by the Indian Govt.

| Date | Measure |
| --- | --- |
| 25/1/2020-13/3/2020 | Health screenings at airports and border crossings |
| 26/2/2020-20/3/2020 | Introduction of quarantine policies: gradually for passengers coming from different countries |
| 26/2/2020-13/3/2020 | Visa restrictions: gradually for different countries |
| 5/3/2020 | Limit public gatherings |
| 11/3/2020 | Border checks |
| 13/3/2020-15/3/2020 | Border closure |
| 16/3/2020 | Limit public gatherings |
| 18/3/2020 | Travel restrictions |
| 20/3/2020 | Testing for COVID-19 |
| 22/3/2020 | Flights suspension |
| 22/3/2020 | Cancellation of Passenger Train Services |
| 24/3/2020 | Suspension of Domestic Airplane Operations |
| 25/3/2020 | 21 days Lockdown of entire Country |
| 25/3/2020 | Cancellation of Passenger Train Services |
| 30/3/2020 | Increase of quarantine/isolation facilities |
| 14/4/2020 | Extension of Lockdown till 3 May 2020 |
| 01/5/2020 | Extension of Lockdown till 17 May 2020 |

## Data Source

We have used Indian COVID-19 data available publicly. The three primary sources of the data are the Ministry of Health and Family Welfare, India (https://www.mohfw.gov.in), https://www.covid19india.org/, and Wikipedia (https://en.wikipedia.org/wiki/2020_coronavirus_pandemic_in_India#Statistics) [^16,17,18^].

### Statistical Models

In this article, we consider the exponential model, the logistic model, and the Susceptible Infectious Susceptible (SIS) model for COVID-19 pandemic prediction at the state level. These models have already been used to predict epidemics like COVID-19 around the world, including China, Ebola outbreak in Bomi, Liberia (2014) [^19,20,21^]. See the appendix for details about the models.

#### Using the models in state-level data

The above three models will provide different prediction perspective for each state. The exponential model-based prediction will give a picture of what could be the cumulative number of infected people in the next 30 days if we do not take any preventive measures. We can consider the forecast from the exponential model as an estimate of the upper bound of the total number of infected people in the next 30 days. The logistic model-based prediction will capture the effect of preventive measures that have already been taken by the respective State Governments as well as the Central Government. The logistic model assumes that the infection rate will slow down in the future with an overall “S” type growth curve. The purpose of the SIS model is to reflect the effect of the major preventive measure like the nation-wide 21-day lockdown from 25 March to 14 April 2020. The lockdown has been extended in two phases, first, till 3 May and then 17 May 2020 with some relaxation [^22,23^]. The SIS model is critically dependent on the infection-rate parameter (β). It is defined as the number of people infected per unit time from an infected person. Note that this parameter is subject to change due to the effect of lockdown and other preventive measures to ensure social distancing. When people are at home, the infection-rate is expected to be on the lower side.

## Study the Effect of Lockdown using daily infection-rate and SIS Model

Kumar et al. [^24^] reported the estimated number of people that a person may *come in contact with within* a day (24 hours) in a rural community in Haryana, India, to be 17. They defined *contact* as having a face-to-face conversation within 3 feet, which may or may not have included physical contact. The estimate of the contact-rate parameter from their paper is 0.70. In practice, only some of all the people who come in *contact* with a COVID-19 infected person may be actually infected by the virus. Note that India has already taken many preventive measures to ensure social distancing. In the current scenario, the infection-rate based on the above study could be an overestimate of its present value. However, despite nationwide lockdown, banks, hospitals, groceries are still open to cater to the essential needs of people. We consider here two approaches to study the effect of lockdown and other preventive measures jointly in each state. *First*, we plot the daily infection-rates for each state. The daily infection-rate (DIR) for a given day is defined as

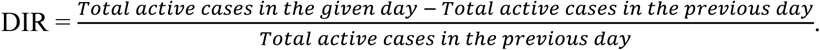

The DIR takes a positive value when we see an increase in active COVID-19 cases from yesterday, the zero value in case of no change in the number of active cases from yesterday, and a negative value when the total number of active cases decreases from the previous day. A DIR value can be more than 1 also, particularly during initial days of infection in a state. For example, when the total number of active cases increases from 5 yesterday to 20 today, then the DIR value is (20-5)/5 = 3. The visual trends in infection rates can provide us whether the COVID-19 situation is under control or not in a specific state. A state where infection-rates are declining for the last few days indicates that the situation is improving. However, a certain jump in infection rates could inform us that there could be cases of COVID-19 that are underreported. We need to search for infected-clusters as quickly as possible. *Second*, using the SIS model, we have considered four predicted line-graphs of active infected patients with different infection-rates. The four different infection-rates, used in the SIS model for prediction are the 25^th^, 50^th^, 75^th^ and 80^th^ percentiles of the observed infection-rates. We also plot the observed active infected patients over time. A declining line-graph of observed active infected patients (red-line) can ensure that measures like lockdown and social distancing are working when all the infected cases are reported/tested. The different predicted lines, using the SIS model, may serve as reference frames to indicate whether the Government needs to enforce the social distancing more stringently. For example, if the current part of the graph of observed active infected patients (red-line) is above the 75 ^th^ percentile line, then there is a major concern for that state. We may need to increase the lockdown period in a state if we do not see the declining graph of observed active infected patients (red-line).

India implemented a nationwide lockdown from 25 March 2020. We first considered the incubation period of novel coronavirus to study the effect of lockdown. The incubation period of an infectious disease is defined as the time between infection and the first appearance of signs and symptoms [^25^]. Using the incubation period, the health researchers can decide on the quarantine periods and halt a potential pandemic without the aid of a vaccine or treatment [^26^]. The estimated median incubation period for COVID-19 is 5.1 days (95% CI: 4.5 to 5.8 days), and 97.5% of those who develop symptoms will do so within 11.5 days (CI: 8.2 to 15.6 days) of infection [^27^]. The WHO recommends that a person with laboratory-confirmed COVID-19 be quarantined for 14 days from the last time they were exposed to the patient [^28^]. Therefore, if a person was infected before the lockdown (25 March 2020), they should not infect others except their family members if that person is entirely inside their house for more than 14 days. WHO also recommends common people to maintain a distance of at least 1 meter from each other in a public place to avoid the COVID-19 infection. The effective implementation of social distancing can stop the spread of the virus from an infected person, even when they are outside for some essential business. However, given a highly dense population in most of India, particularly in cities, it may not always be possible to maintain adequate social distance.

### State-wise Analysis and Prediction Report

In this section, we depend on inputs from the exponential, logistic, and SIS models along with daily infection-rates for each state. Remembering the word of the famous statistician George Box “All models are wrong, but some are useful,” we interpret the results from different models jointly. We consider different states with at least 300 cumulative infected cases. For each state, we present four graphs. We have used the state-level data until 1 May 2020. The first and second graphs are based on the logistic and the exponential models, respectively, with the next 30 days predictions. The third graph is the plot of daily infection-rate for a state. Finally, the fourth graph is showing the growth of the active infected patients using SIS model prediction *(“pred”)* along with the observed active infected patients. We do not show the next 30 days predictions (which are very high values) using the SIS model to ensure the distinguishability of the different line-graphs. Table 2 represents the 30-days prediction of the cumulative infected number of people for each state using the logistic model, the exponential model, and a data-driven combination of the two (see below for details). The corresponding measures of goodness of fit (R-square and Deviance) are presented in Table 3 within the Appendix.

**Table 2:**
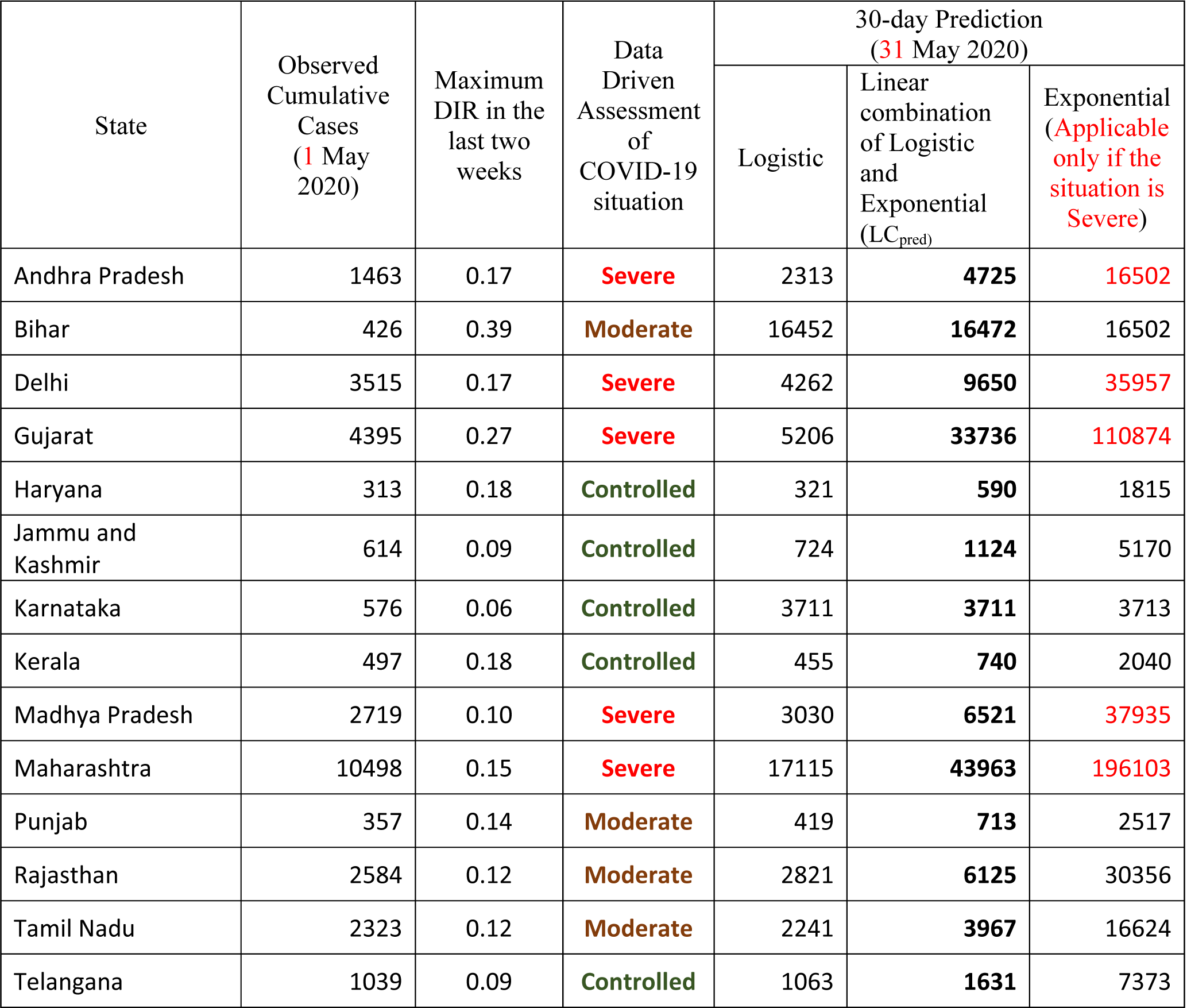
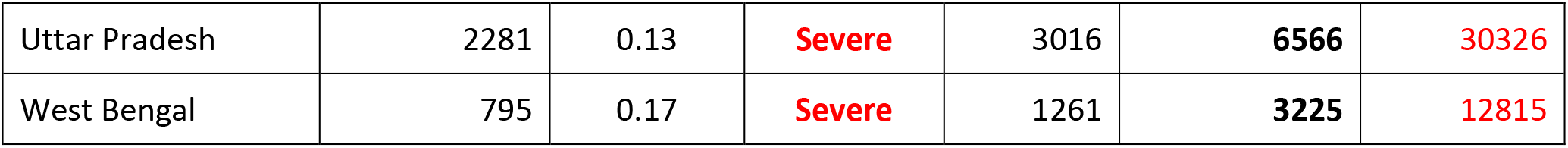
Data-driven assessment and 30--day prediction using the logistic and exponential models and their linear combination.

**Table 3.** Goodness of fit measures for the logistic and exponential models.

| State | Logistic |  | Exponential |  |
| --- | --- | --- | --- | --- |
|  | R-square | Deviance | R-square | Deviance |
| Andhra Pradesh | 0.99 | 70932.20 | 0.99 | 169609.12 |
| Bihar | 0.99 | 8244.24 | 0.99 | 8243.97 |
| Delhi | 0.99 | 563263.44 | 0.97 | 1979522.61 |
| Gujarat | 1.00 | 222500.23 | 0.98 | 1869895.38 |
| Haryana | 0.99 | 10970.46 | 0.93 | 49223.34 |
| Jammu and Kashmir | 0.99 | 14326.21 | 0.97 | 62115.67 |
| Karnataka | 0.97 | 61993.09 | 0.97 | 62001.89 |
| Kerala | 0.99 | 22603.01 | 0.91 | 287919.78 |
| Madhya Pradesh | 0.99 | 330231.58 | 0.96 | 1406825.35 |
| Maharashtra | 1.00 | 437647.58 | 0.99 | 4452503.81 |
| Punjab | 0.99 | 4082.03 | 0.97 | 24317.88 |
| Rajasthan | 1.00 | 58694.90 | 0.97 | 1379677.74 |
| Tamil Nadu | 0.99 | 357650.72 | 0.95 | 1740108.65 |
| Telangana | 1.00 | 29424.88 | 0.92 | 797659.71 |
| Uttar Pradesh | 1.00 | 33738.33 | 0.98 | 541768.49 |
| West Bengal | 1.00 | 4001.97 | 0.99 | 32894.14 |

#### Maharashtra

The situation in Maharashtra is currently very severe with respect to the active number of cases. As of 1 May 2020, the total number of infected cases is 10498. The logistic model indicates that in another 30 days from now, the state can observe around 17100 cumulative infected cases. The daily infection-rates for this state are between 0.03 and 0.15 in the last two weeks, and it was more than 0.4 for two days at the beginning of April. Note that, for Maharashtra, the lower DIR values of 0.03 may not indicate a good sign since the total number of *active infected cases* is above 8000. Thus, a DIR value of 0.03 for a day implies 8000 x 0.03 = 240 new infected cases. The line-graphs from the SIS model are alarming as the observed active infected patients (red-line, 4^th^ panel) line is far above the predicted line with estimated infection-rate at the 80^th^ percentile (β = 0.22). It is apparent from the graphs that even after 30 days of lockdown, Maharashtra has not seen any decline in the number of active cases. This may also indicate that there could be a large number of people who are in the community without knowing that they are carrying the virus. The state can be considered in stage 3.

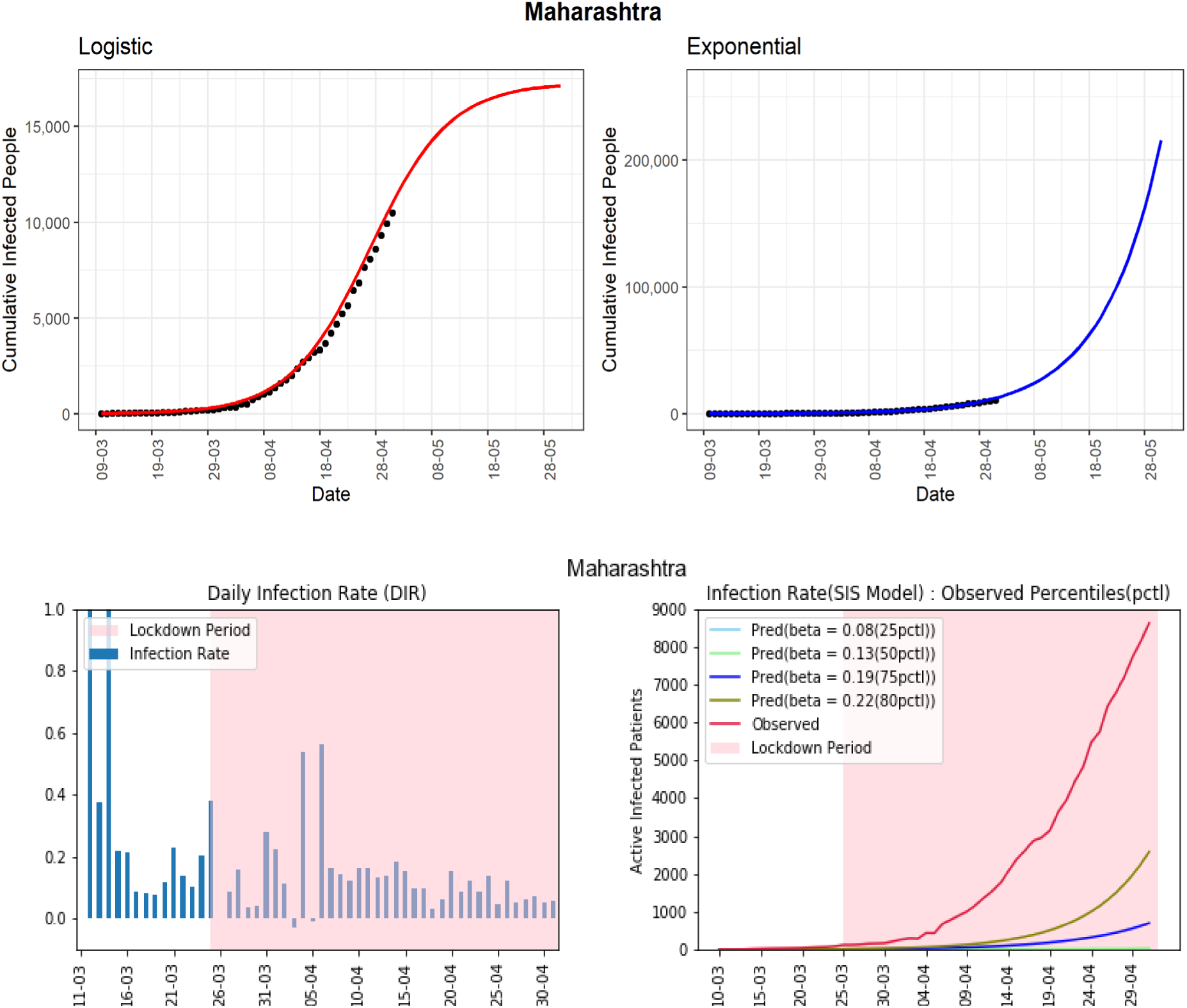

#### Delhi

Delhi, being a high population-density state, has already observed 3515 confirmed COVID-19 cases. Based on the logistic model, the predicted number of cumulative infected cases could reach around 4200 in the next 30 days. The daily infection-rate (DIR) has not seen a downward trend in the past few days. The line-graph (red-line, 4^th^ panel) of observed active infected patients was showing a downward trend during 20-23 April. However, the same graph has picked up exponential growth in the last few days. This an important observation that indicates us why we need a continuous downward trend of active cases for at least 14 days, a slight relaxation may put a state in the same severe condition where it was earlier. The observed infection-rate is currently fluctuating between −0.06 and 0.17 in the last two weeks. The occasional high infection-rate may suggest that there could be many people who are in the community without knowing that they are already infected with the COVID-19. The state could be heading to community spreading of COVID-19 (stage 3).

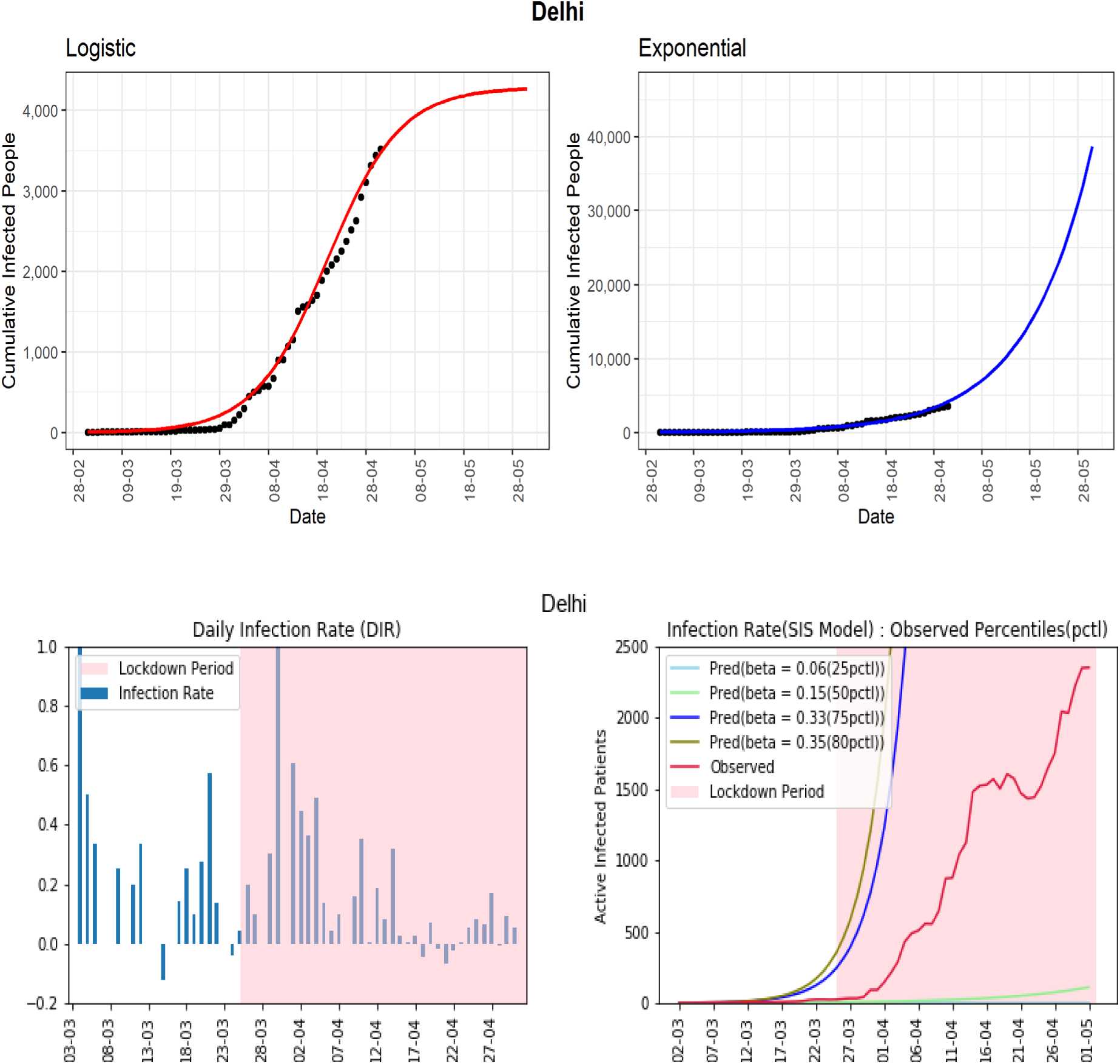

#### Tamil Nadu

The cumulative infected cases in Tamil Nadu is 2323. The state has observed a high daily infected-rate of more than 0.7 in some days in March. Tamil Nadu is one of the states where the effect of lockdown is visibly seen from the declining daily infected-rates from the beginning to the end of April. However, there is again an increasing trend in the last three days in DIR. The DIRs are in between −0.13 and 0.12 over the previous two weeks. The later part of the line-graph (red-line, 4^th^ panel) of observed active infected patients is showing a decreasing trend first, then again an increasing trend. The preventive measures need to be maintained to bring down the active cases as well as to stop a new infection in this state.

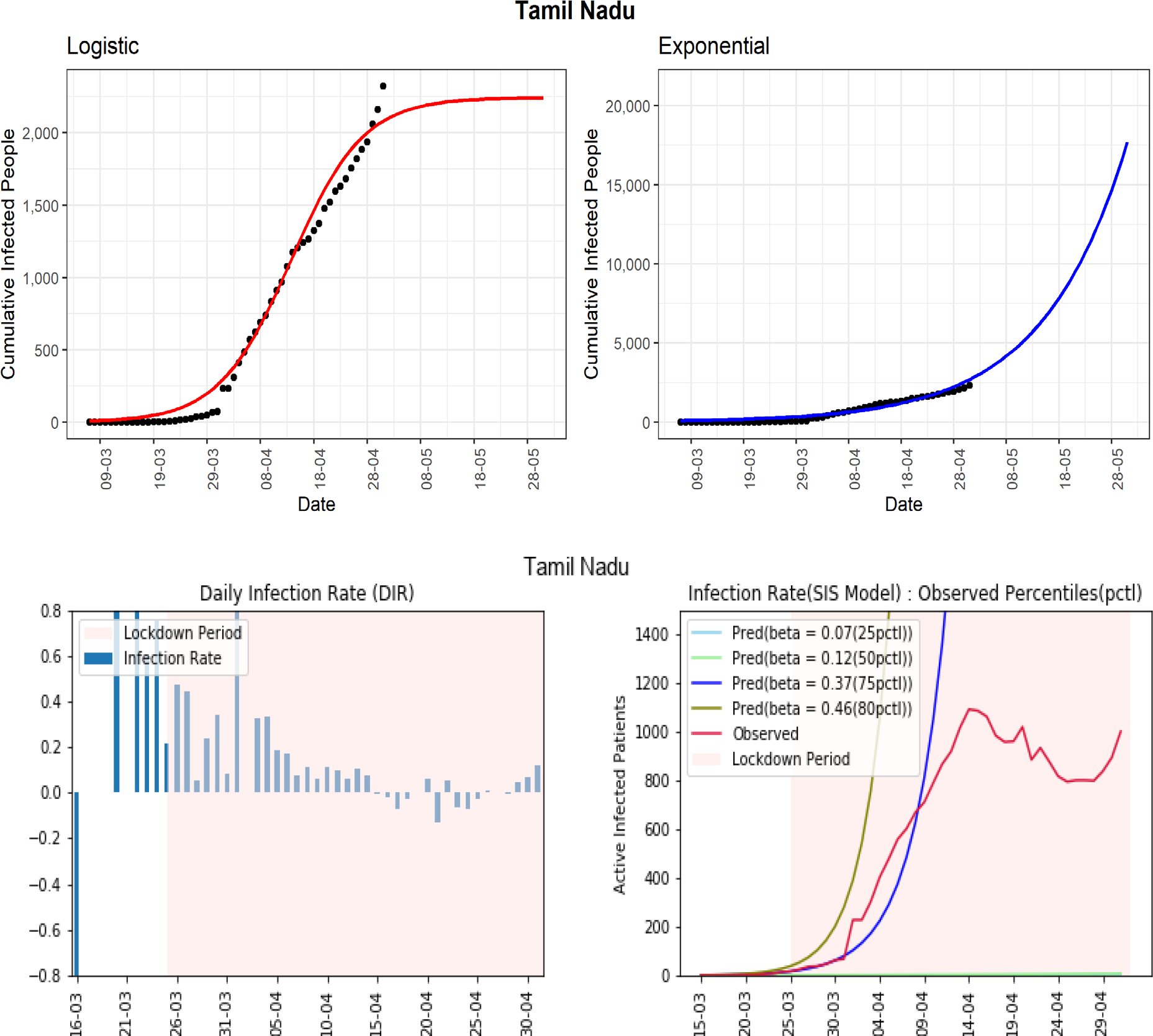

#### Madhya Pradesh

The state currently has 2719 cumulative COVID-19 cases. In the later part of the lockdown, after 10 April, the state observed a few days with infection-rate more than 0.4. Till now, there is no sight of a declining trend in the daily infected-rates. The same type of conclusion can be drawn from the line-graphs of the SIS model. The line-graph (red-line, 4^th^ panel) of observed active infected patients is in between the lines corresponding to 50^th^ – 75^th^ percentiles line-graphs. The same line-graph is maintaining an exponential growth after 10^th^ April. Notice that, for Madhya Pradesh, the 50^th^ percentile of observed infection-rates is 0.14, which is higher than the 50^th^ percentile of some other states. The high growth of active cases in the latter part of the lockdown is a major concern for this state. It could be a signal of a community spread of the COVID-19.

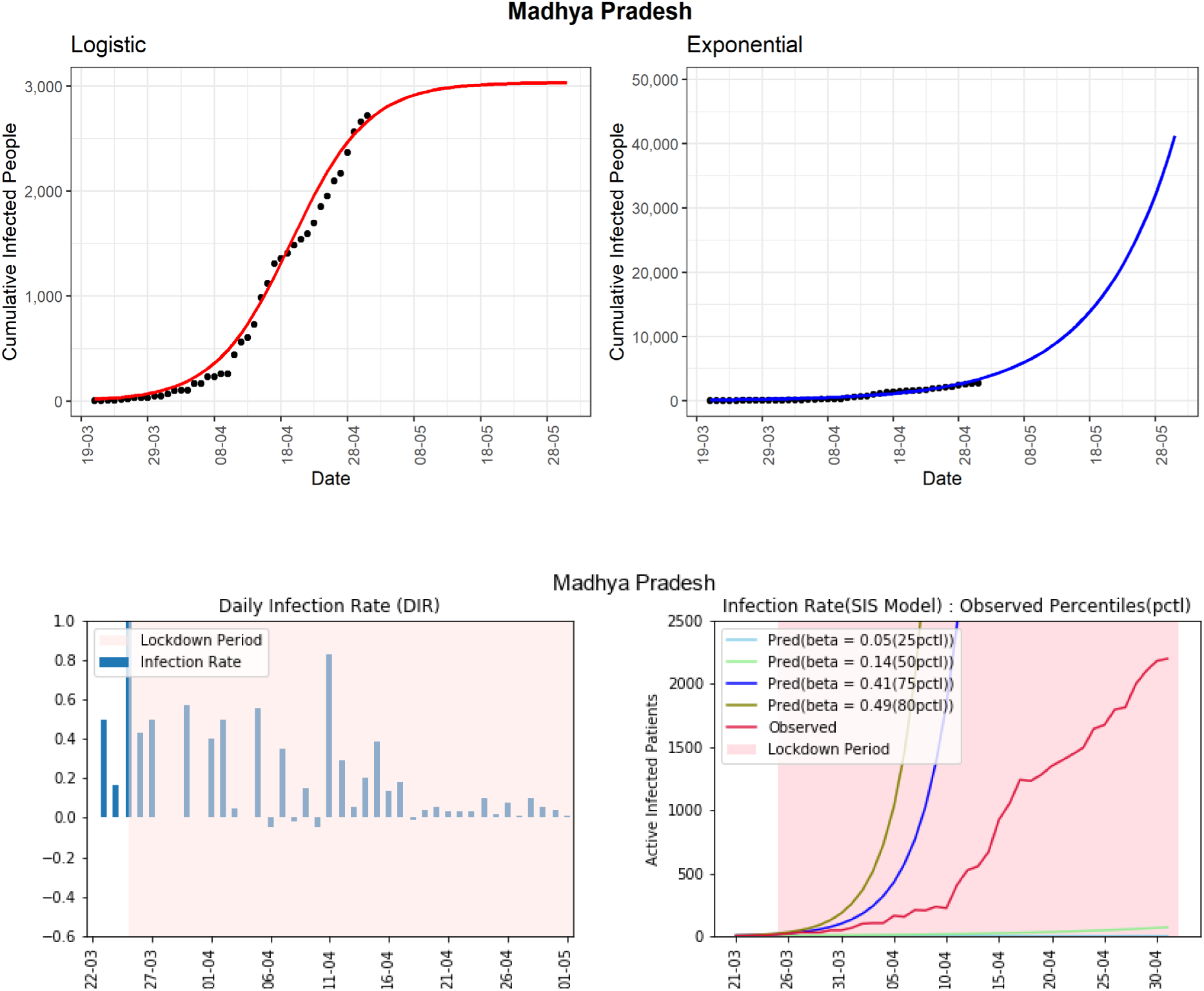

#### Rajasthan

The western state of India, Rajasthan, reported 2584 cumulative infected COVID-19 cases. The logistic model indicates that in another 30 days from now, the state can observe around 2800 cumulative infected cases. The state has seen a declining trend in the daily infected rates during the last part of April. The line-graph (red-line, 4^th^ panel) of observed active infected patients is increasing and is in between the curves of 50^th^-75^th^ percentiles of observed infection-rates (0.14-0.27) using the SIS model. In the last two weeks, the DIRs for Rajasthan are fluctuating between −0.05 and 0.12. The active cases in this state have not increased too much in the latter part of April. An increase in recovery cases is one of the reasons. However, the current COVID-19 situation in the state is not controlled yet.

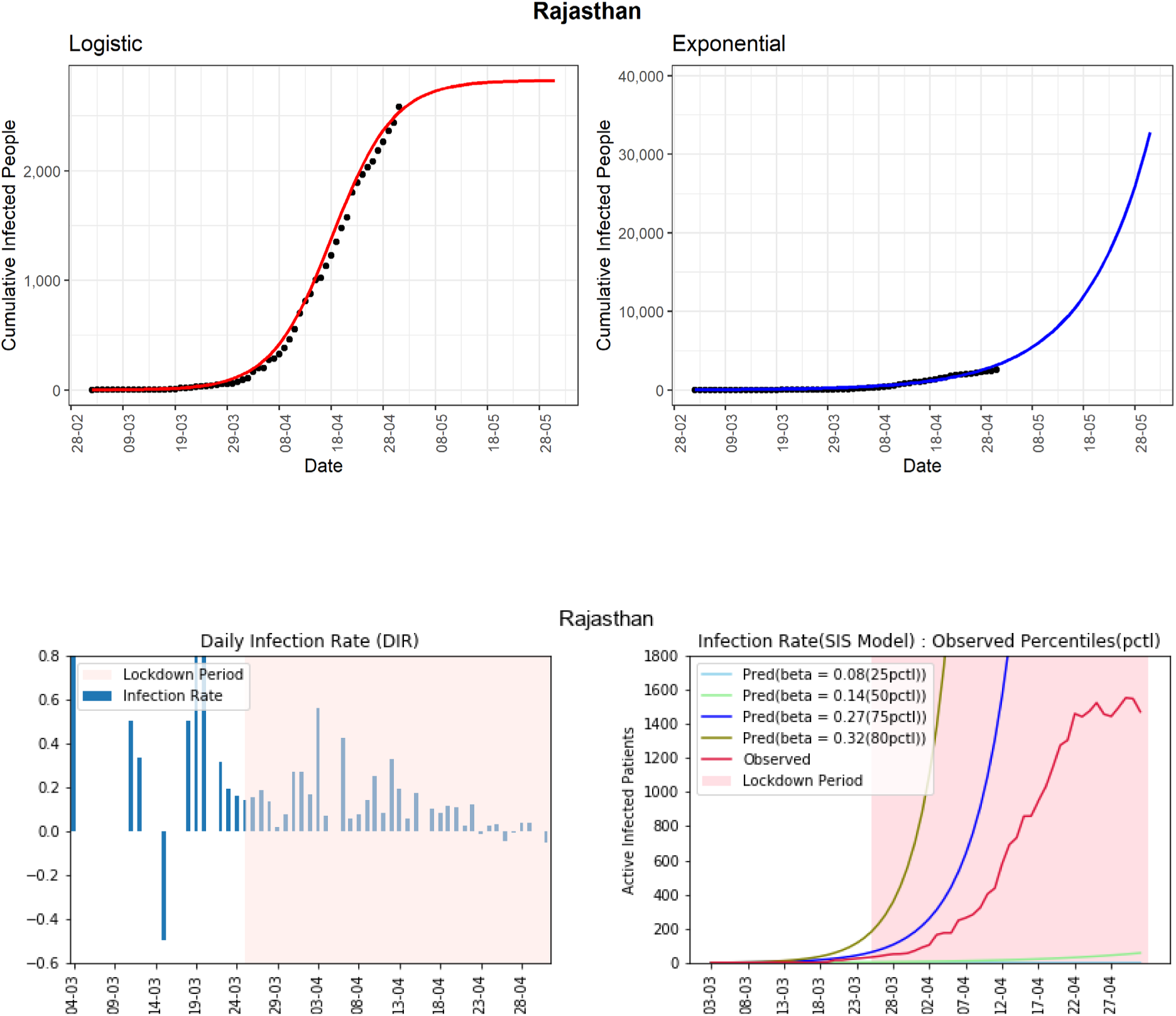

#### Gujarat

The state is currently experiencing exponential growth with 4395 as the cumulative number of COVID-19 cases. Using the logistic model, the cumulative infected cases could reach around 5206 in the next 30 days. There is an apparent stable rather than a declining trend in the daily infection-rates in the last few days. The DIRs are in the range of 0.03 to 0.27 in the last two weeks, which are on the higher side. The line-graph (red-line, 4^th^ panel) of observed active infected patients is close to the curve of the estimated 75^th^ percentile of observed infection-rate (β = 0.26). Surprisingly, in the latter part of the lockdown, the red-line is experiencing exponential growth. This state needs immediate intervention to implement all the preventive measures already taken by the Government strictly.

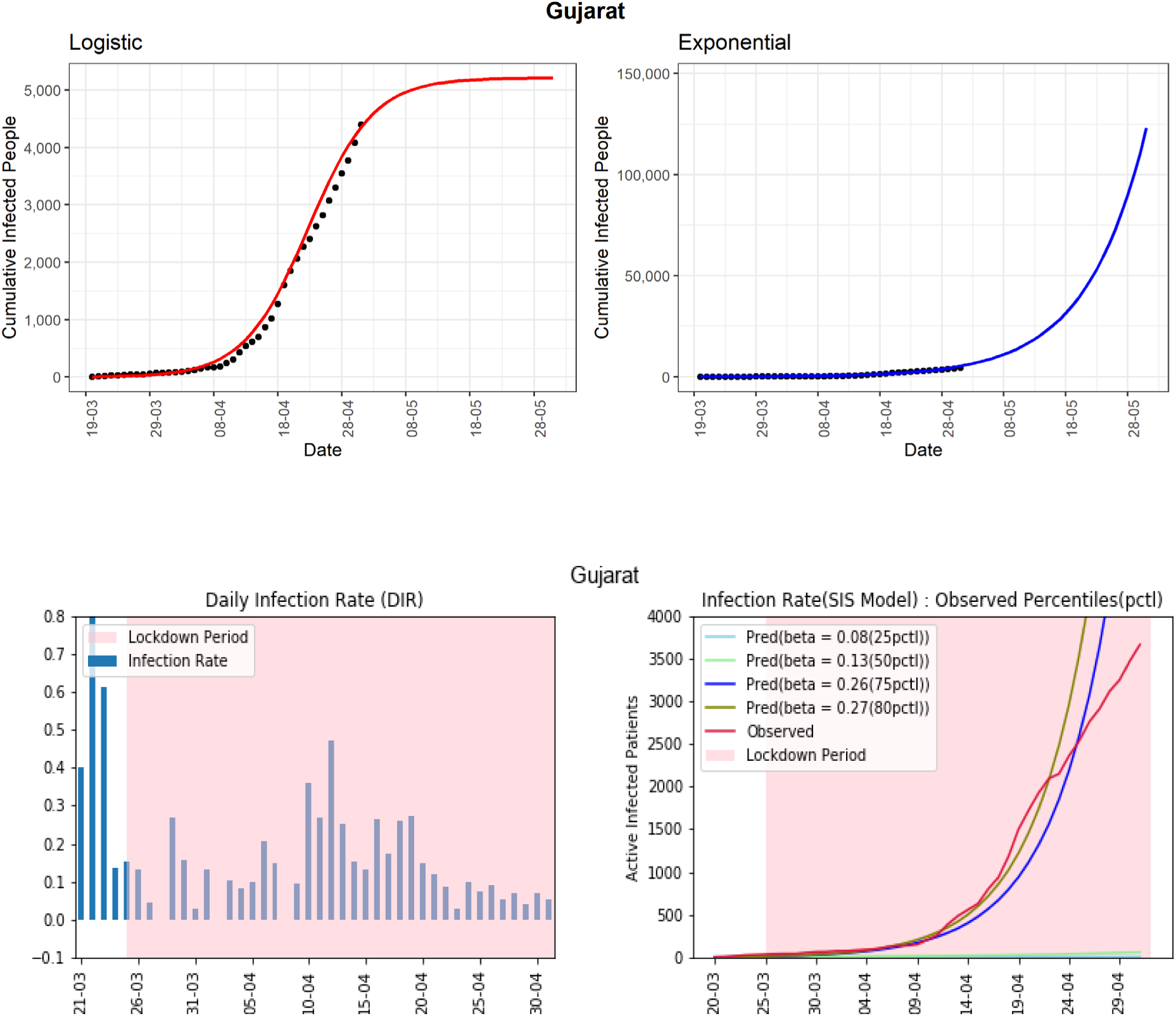

#### Uttar Pradesh

This northern state of India has experienced 2281 cumulative COVID-19 cases. Using the logistic model, the predicted number of cumulative confirmed cases could be around 3000 in the next 30 days. The line-graph (red-line, 4^th^ panel) of observed active infected patients is in between the line-graphs of the 50^th^ and 75^th^ percentiles of observed infection-rates (β = (0.12, 0.23)). The daily infection-rate is in the range of −0.02 to 0.13 without a moderate decreasing trend in the last two weeks. The overall growth of active cases is still exponential, which is a major concern for the state. There could be many unreported cases in the state. In the absence of preventive measures, unreported cases can contribute to spreading the virus in the community.

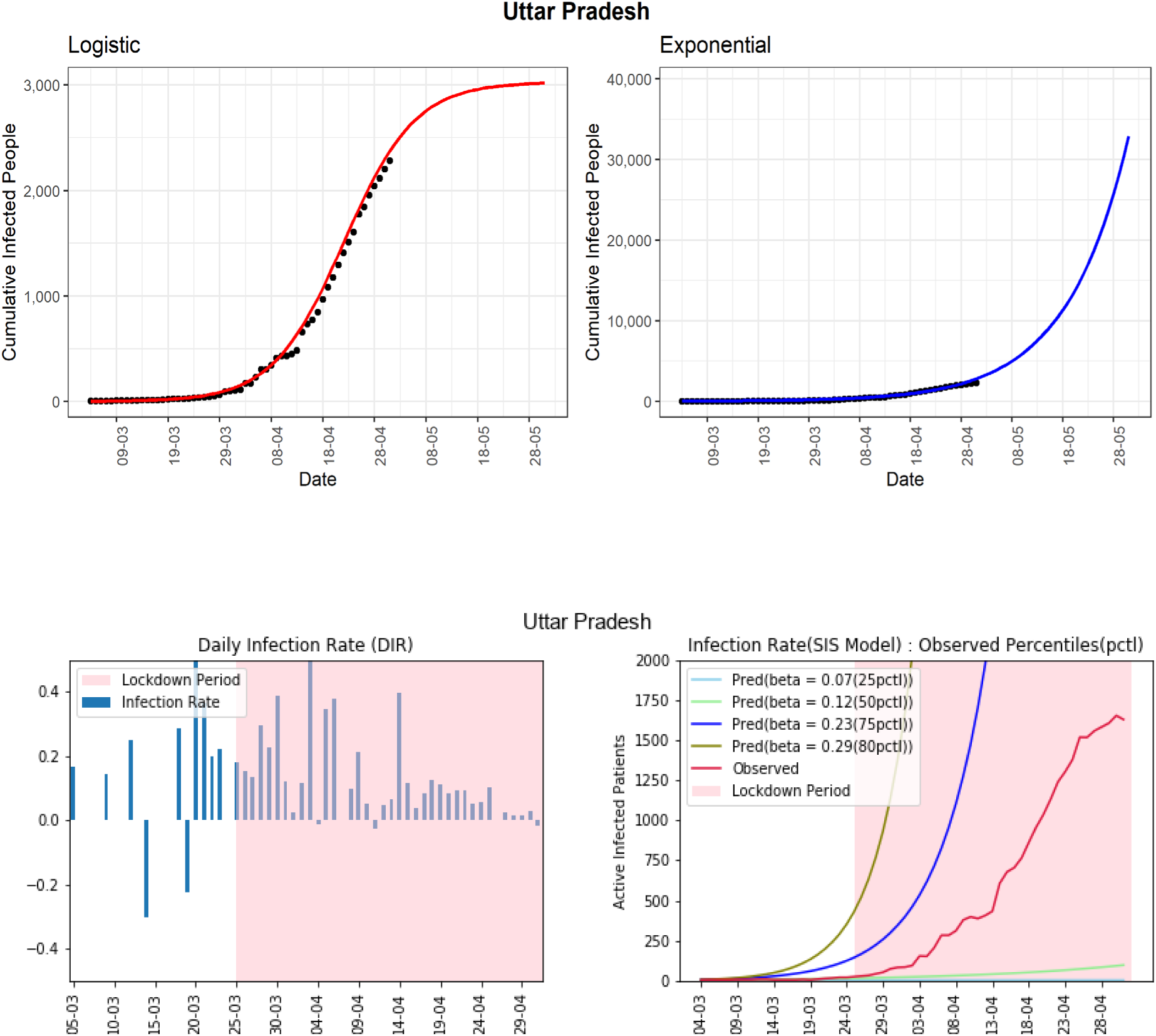

#### Telangana

The southern Indian state of Telangana has reported 1039 cumulative infected cases till now. The logistic model predicts that the number of cases for the state will be around 1063 in the next 30 days. In the fourth graph, the line-graph (red-line, 4^th^ panel) shows that the active number of cases has continuously remained below the curve of the 75^th^ percentile of the observed infection rate (β = 0.25). From 23 April onwards, there is a visible downward trend in the same line graph. This evidence is also supported by a clear decreasing trend in the daily infection rate for more than two weeks. The state is going in the right direction to control the COVID-19 pandemic. However, preventive measures need to be in place to see long term success again the virus.

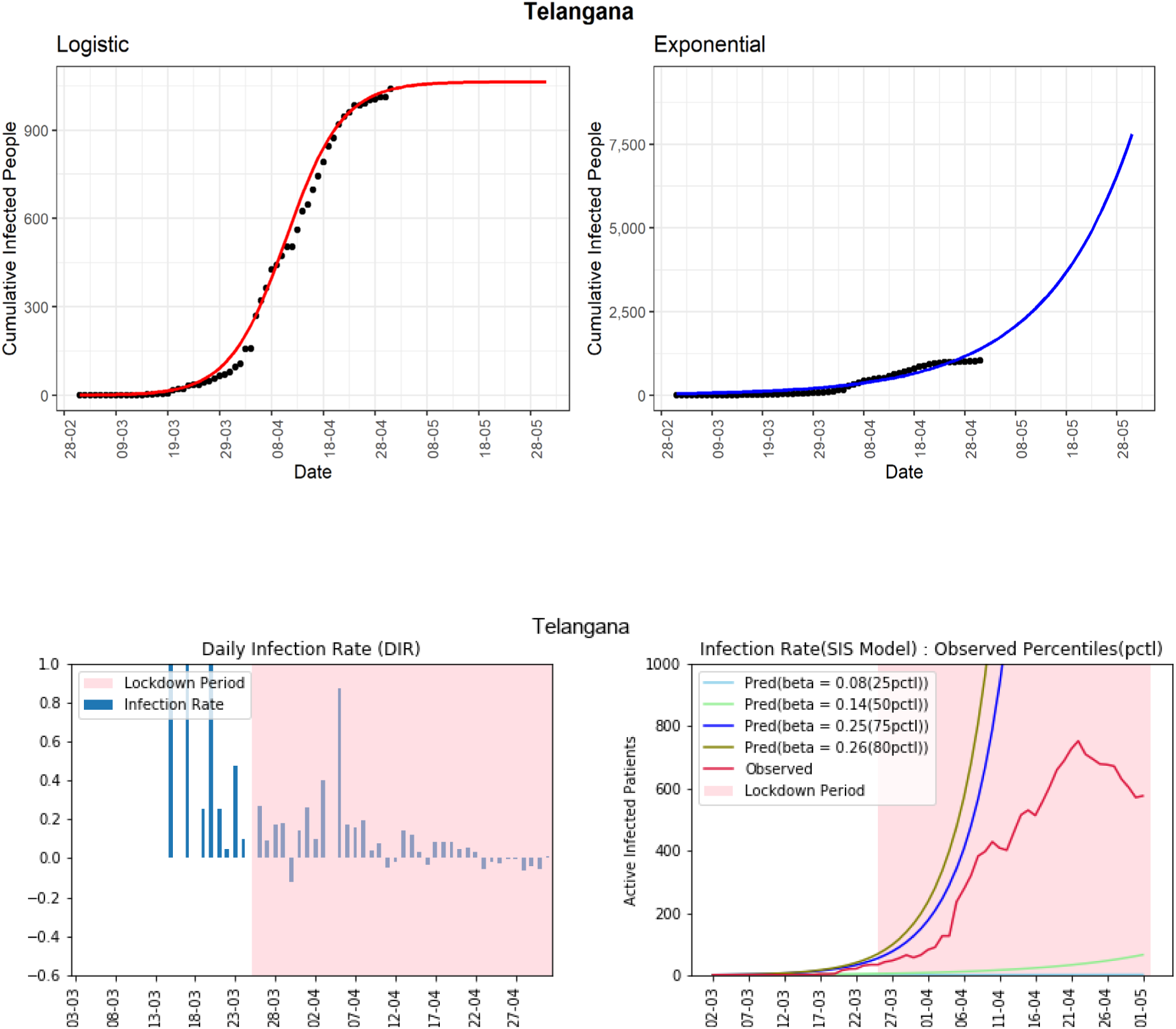

#### Andhra Pradesh

The state has observed 1463 confirmed cumulative infected cases so far. The line-graph (red-line, 4^th^ panel) shows that the number of active cases is now below and close to the curve of the 75^th^ percentile of the observed daily (β = 0.23) infection rate. The logistic model is showing the maximum number of cumulative infected people will be around 2313 in the next 30 days. Despite showing good progress in mid-April, the state is again showing an exponential type growth rate. This state has seen daily infection rates in between −0.04 to 0.17 during the last two weeks. The state start has shown few short declining trends but without a long-term decline trend in the DIR values. It could be due to many unreported infected cases in the community that is spreading the virus.

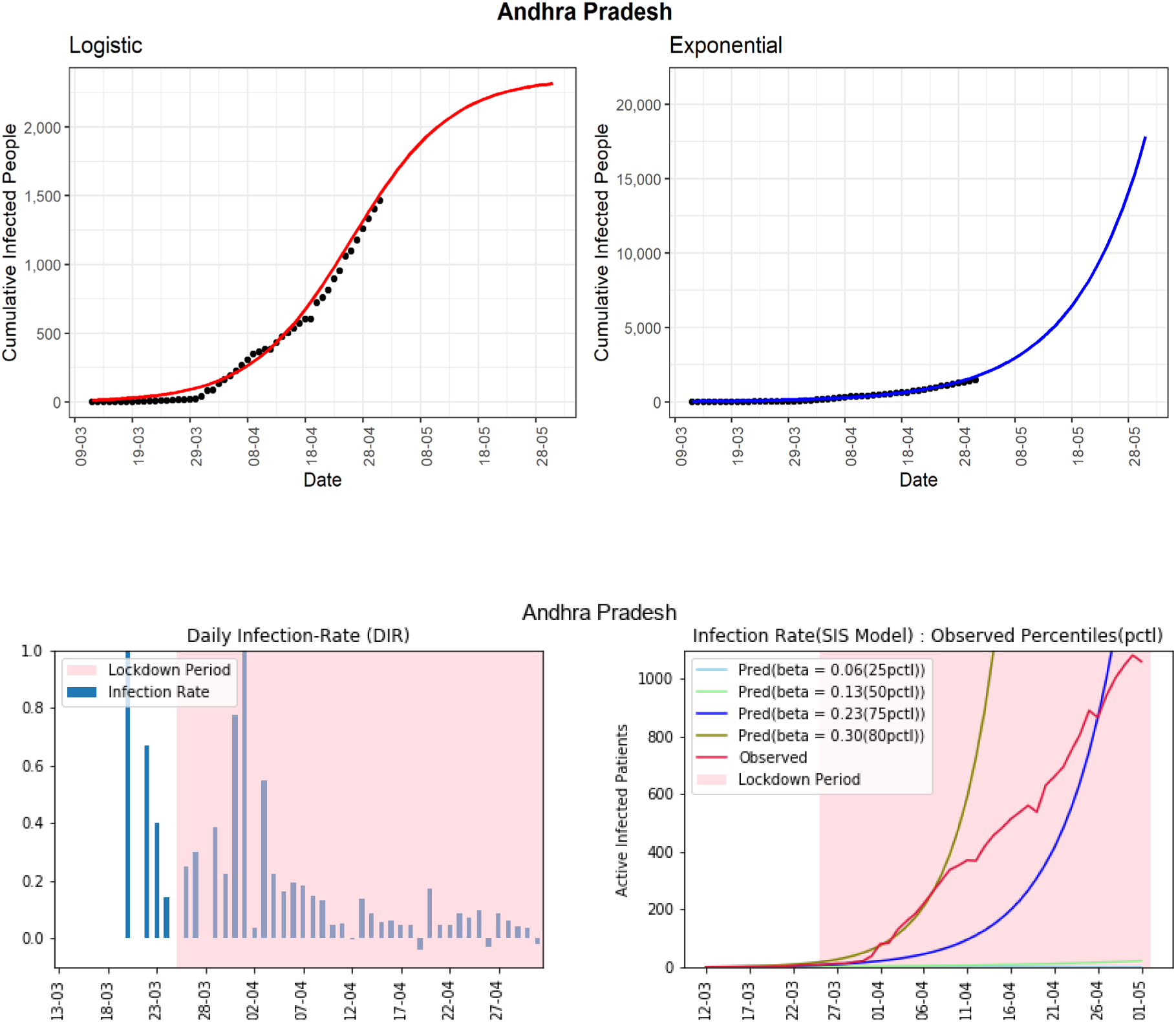

#### Kerala

The southern state of Kerala is one of the few states of India, where the effect of the lockdown is observed strongly. The state reported the first COVID-19 case in India. However, Kerala has been able to control the spread of the virus to a large extent to date. The cumulative number of cases reported until now is 497. It is a state where the line-graph (red-line, 4^th^ panel) of observed active infected patients is going down, which shows that the lockdown and other preventive measures have been effective for this state. The daily infection-rate has declined steadily from positive to negative values. However, some spikes in the DIR values can be noticed in the last few days. It can be expected that with the present scenario of the extended lockdown, the number of active cases will be few at the end of May.

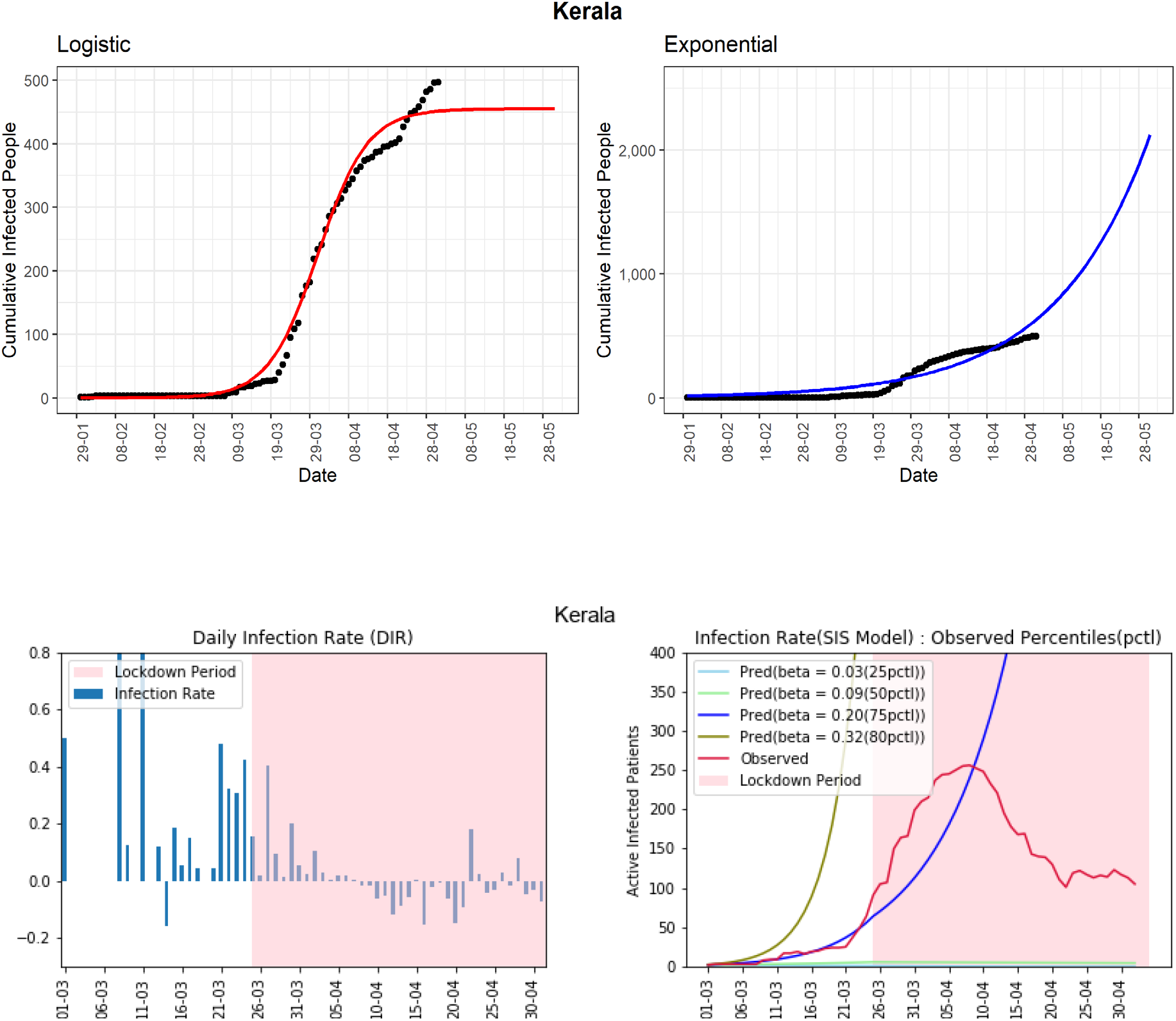

#### Karnataka

The state has managed to restrict the cumulative infected cases to 576 till now. The line-graph (red-line, 4^th^ panel) of observed active infected patients is now below the line-graph of the 75 ^th^ percentile of the observed infection rate (β = 0.18). Compared to other states, the 75^th^ percentile infection-rate is on the lower side. We can observe the ups and downs of the daily infected-rate with upper bound as 0.2 from early April. This state has seen daily infection rates between −0.04 to 0.06 during the last two weeks. However, the preventive measure needs to be maintained to control the spread of the virus.

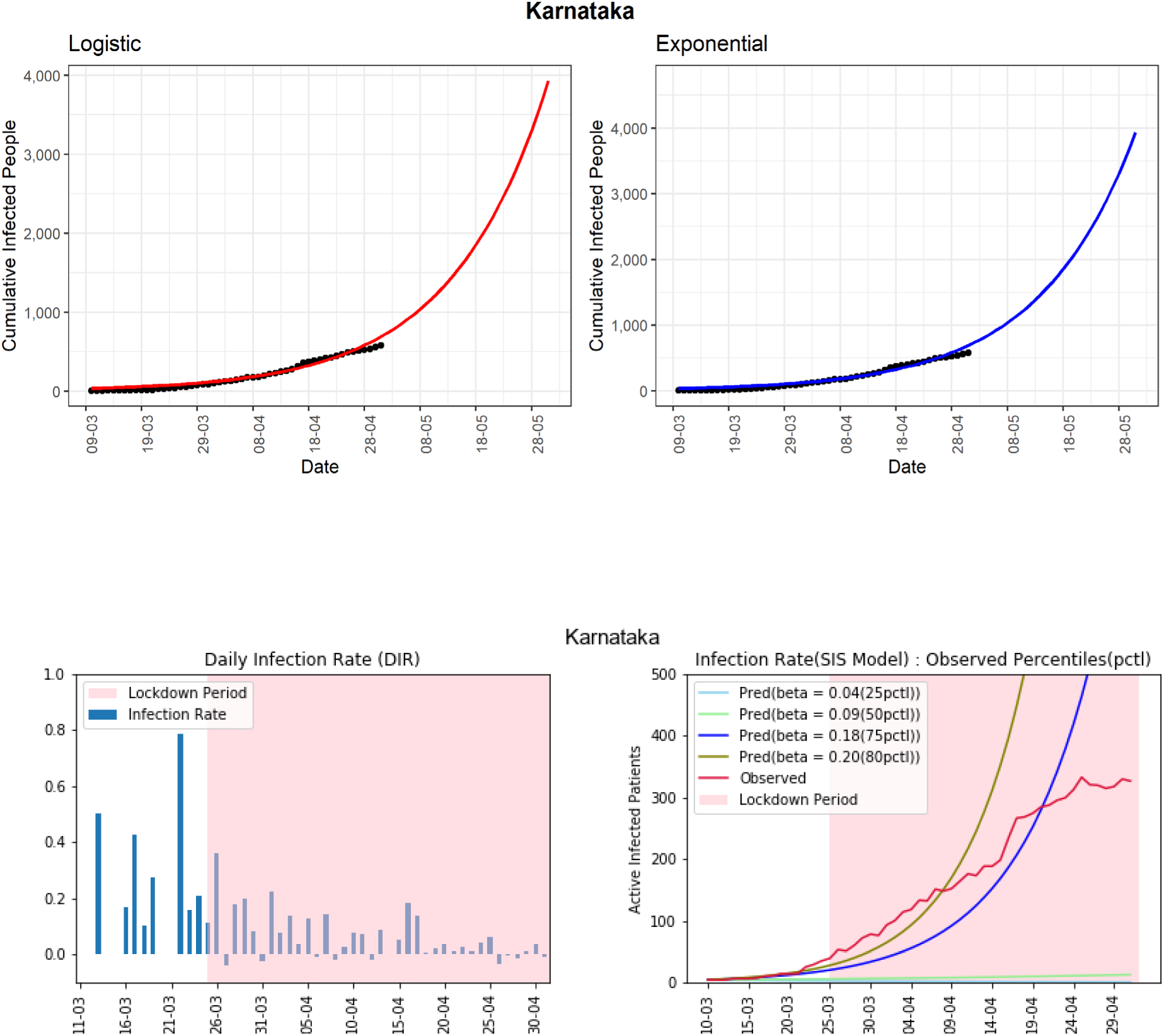

#### Jammu and Kashmir

The northernmost state of Jammu and Kashmir has seen 614 cumulative infected cases so far. The line-graph (red-line, 4^th^ panel) of observed active infected patients has been far below the 75^th^ percentile of the observed daily infected rate (β = 0.35). From 9 April, the daily infection-rate is apparently decreasing. There are some spikes in DIR values occasionally. It could be due to many unreported cases, which are allowing infection to spread even during the lockdown period. The daily infection-rate is in the range of −0.02 to 0.09 in the last two weeks.

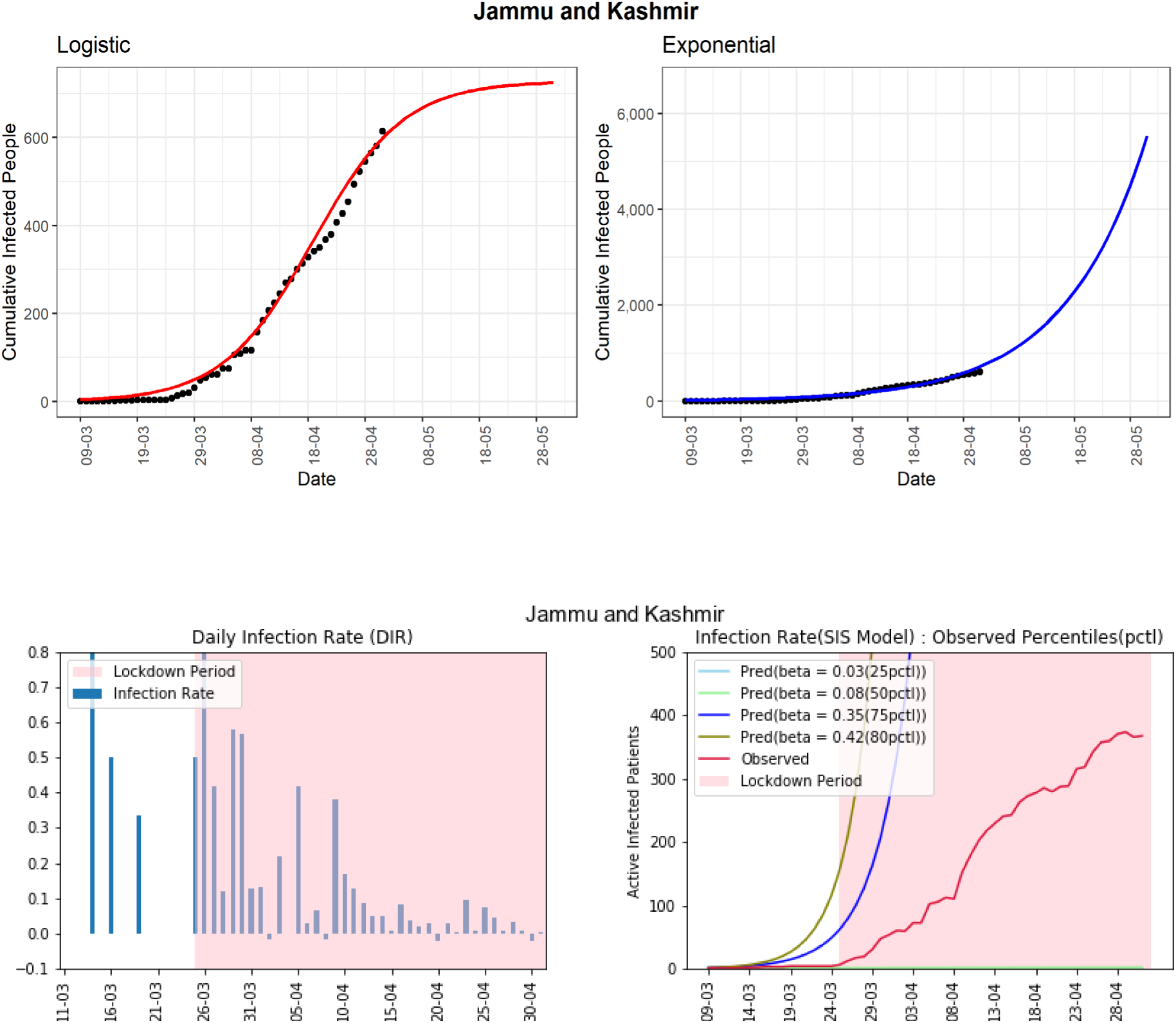

#### West Bengal

The state of West Bengal is standing at 795 cumulative infected cases as of now. The daily infection-rate does not show any trend of slowing down in recent times. Based on the logistic model, the predicted cumulative infected cases can be around 1261 in the next 30 days. The line-graph (red-line, 4^th^ panel) of observed active infected patients is above the line-graph of the 75 ^th^ percentile of the infection rate (β = 0.21) graph. The DIRs are between 0.03 and 0.17 in the last two weeks. All the three graphs, cumulative infected cased based logistic and exponential graphs, and active case-based line-graph (red-line, 4^th^ panel) are all showing exponential type growth rates. Strict implementation of preventive measures is needed to control the spread of the COVID-19 in the state.

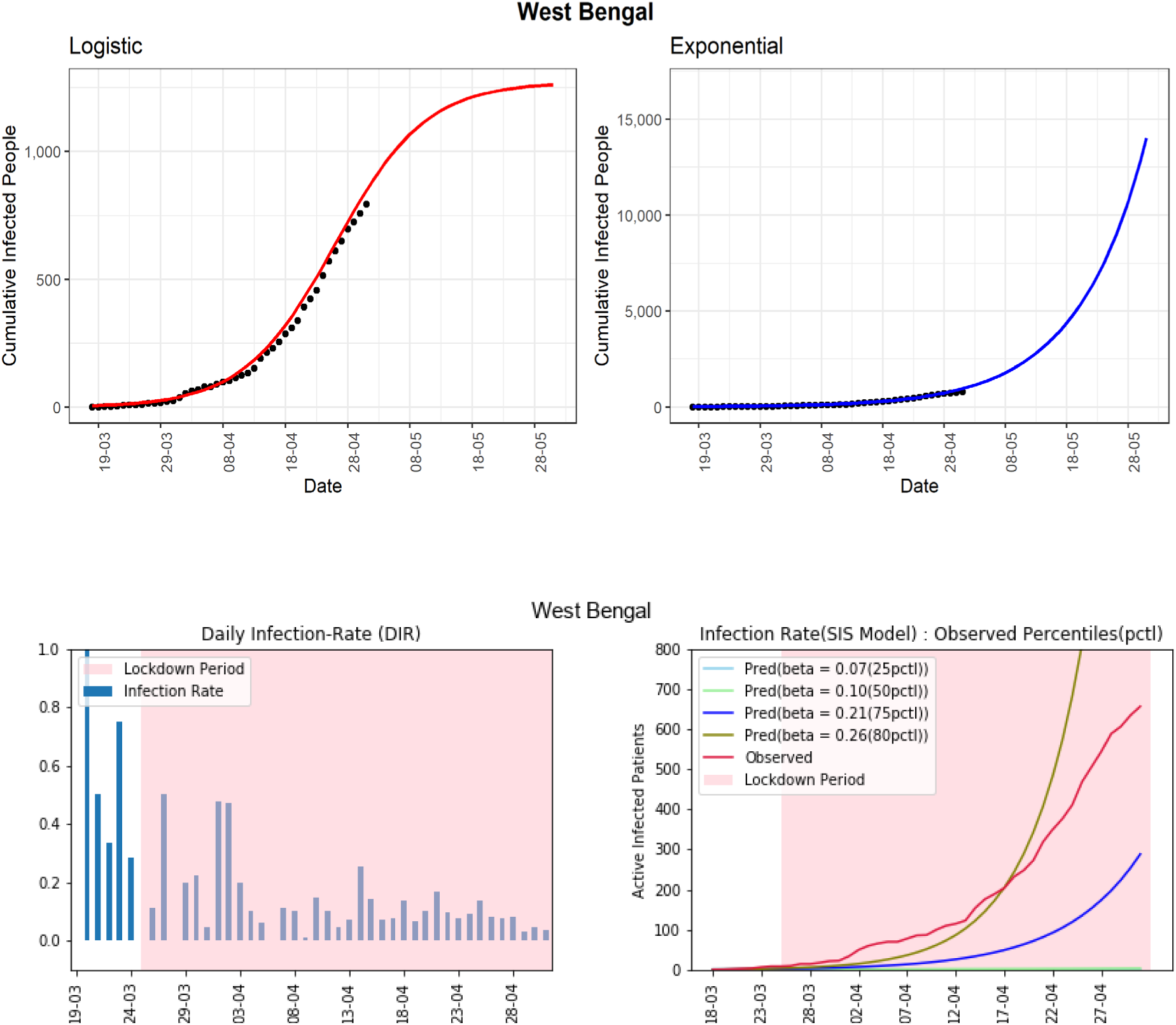

#### Haryana

The state of Haryana observed 313 cumulative infected COVID-19 cases so far. It has reported a very low rate of infection-rate in the latter part of the lockdown except for the last reported day. In the 4^th^ panel, the line-graph (red-line) of observed active infected patients is now far below the line-graph of the 50^th^ percentiles (β = 0.15) and showing a decreasing trend in the latter part. The DIRs are between −0.28 and 0.18 in the last two weeks. Under the assumption that there are not too much-unreported cases, the situation in Haryana seems to be under control.

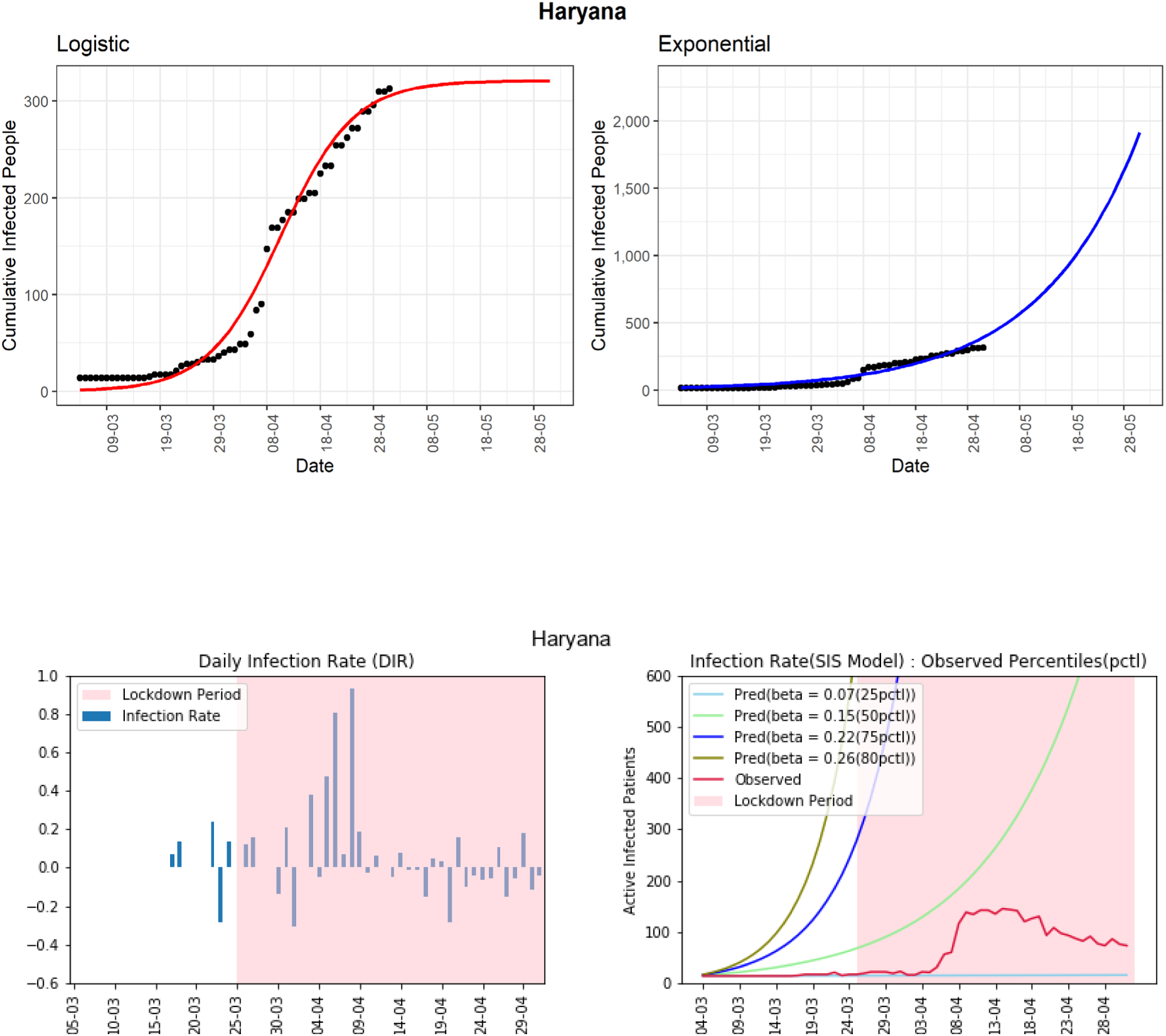

#### Punjab

The state of Punjab has reported 357 cumulative infected cases till now. Based on the logistic model, the predicted cumulative confirmed cases could be around 419 in the next 30 days. The line-graph (red-line) of observed active infected patients is in between the curves of the estimated 75^th^ and 80^th^ percentiles of observed infection-rate (β = 0.15, 0.28). The daily infection-rates are between −0.05 and 0.14 in the last two weeks, which is good given the low number of active infected cases in the state.

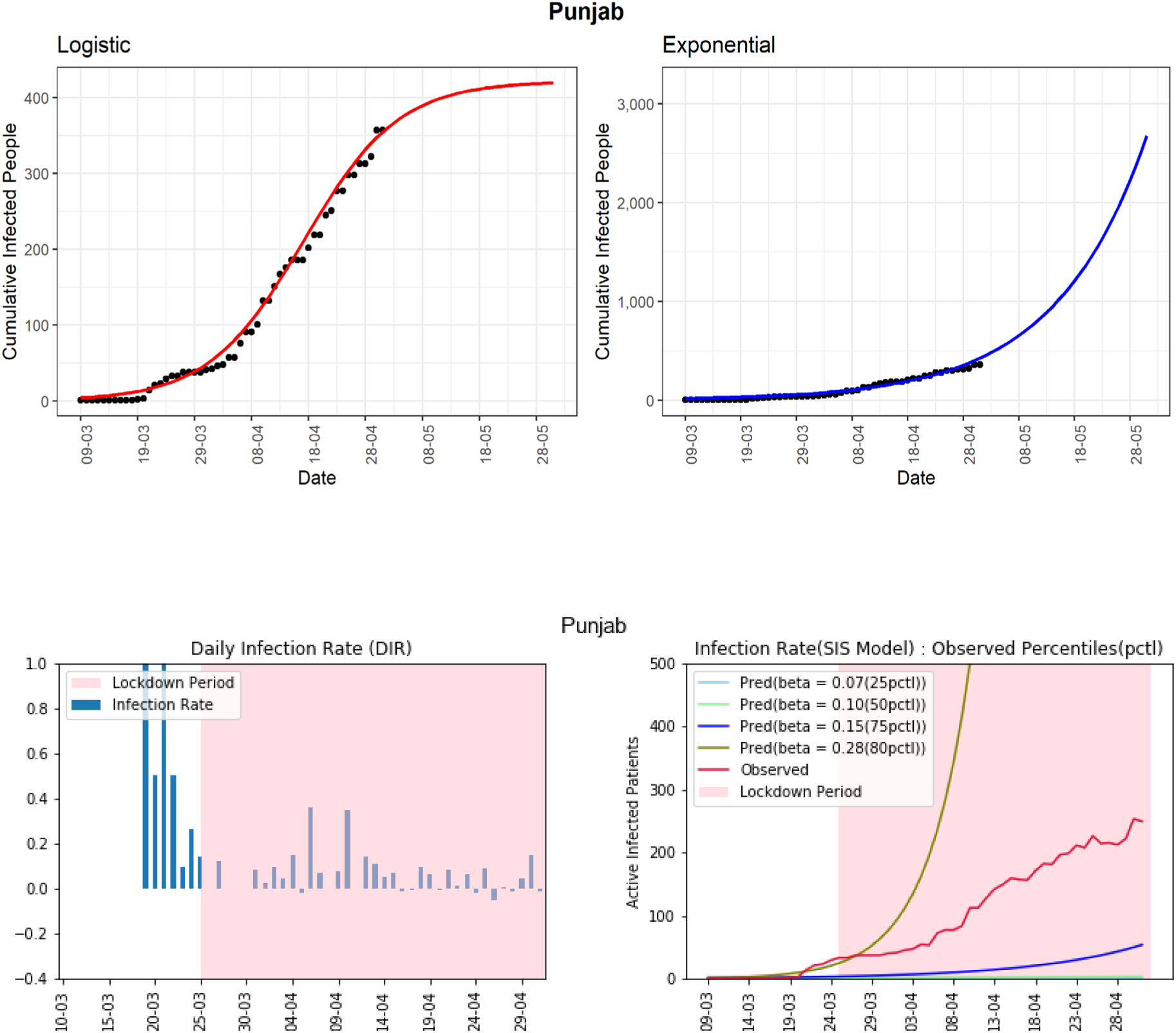

#### Bihar

The state has reported 426 cumulative infected cases until now. Based on the logistic model, Bihar could see 16452 total infected cases in the next 30 days. It may be an overestimate. However, the DIRs show no sign to decline in the last two weeks, with the highest reported value of 0.39. It may indicate many unreported cases in the state. However, the cumulative infected cases are still low for this state. Effective implementation of preventive measures is needed for the state.

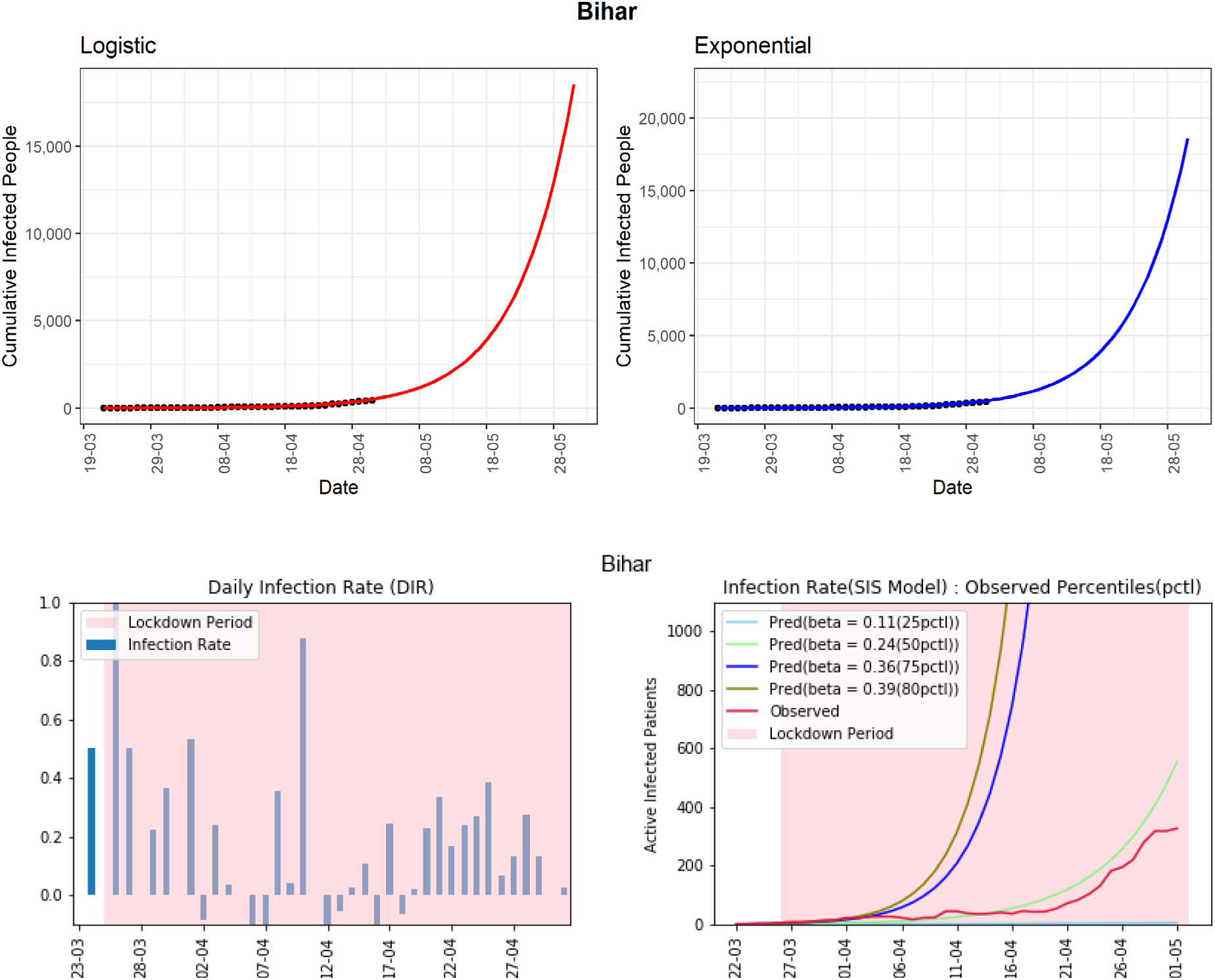

### Joint Interpretation of Results from all Models

We consider a data-driven assessment of the COVID-19 situation based on the growth of active cases in recent times (red-line, 4^th^ panel in each state plot) along with the DIR values for each state (see Table 2). We label a state as *severe* if we observe a non-decreasing trend in DIR values over the last two weeks and a near exponential growth in active infected cases; as *moderate* if we observe an almost decreasing trend in DIR values over the last two weeks and neither increasing nor decreasing growth in active infected cases; and as *controlled* if we observe a decreasing trend in the last two weeks’ DIR values and a decreasing growth in active infected cases. It can be noticed that the logistic model is under-predicting the next 30-day prediction, whereas the exponential model is over-predicting the same. As we argued earlier, despite nationwide lockdown, people are out of the home for essential businesses, which can contribute to spreading the virus. The maximum value of DIR in the last two weeks can capture how severely the COVID-19 is spreading in recent times. Note that the DIR value of 0.10 cannot be interpreted in a similar way for two different states, say, with 500 and 5000 active cases, respectively. For the first state, we see 500 x 0.10 = 50 new cases and for the second state, we observe 5000 x 0.10 = 500 new cases. In an attempt to capture these various subtleties in a realistic prediction, we propose a linear combination (LC_pred_) of the logistic and the exponential predictions using the maximum value of DIR over the last two weeks (DIR_max_) as a weighting coefficient (tuning parameter) as follows:

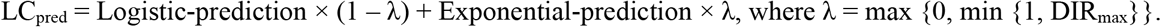

Such a choice of the tuning parameter *λ* makes the LC_pred_ equal to the logistic prediction when DIR_max_ is negative with λ = 0. On the other hand, the LC_pred_ is equal to the exponential prediction when DIR_max_ is more than 1 with λ = 1. When DIR_max_ is in between 0 and 1, the LC_pred_ is a combination of the predictions from the logistic and the exponential models. Given the situation in entire India, we recommend LC_pred_ to be used for assessment purposes for each state.

## Discussion

India, a country of 1.3 billion people, had reported 17615 confirmed COVID-19 cases after 80 days (from 30 January 2020) from the first reported case in Kerala [^29^]. In a similar duration from the first case, the USA reported more than 400,000; both Spain and Italy reported more than 150,000 confirmed COVID-19 cases. To gain some more perspective, note that, the USA has around one-fourth of the Indian population. Therefore, according to the reported data so far, India seems to have managed the COVID-19 pandemic better compared to many other countries. One can argue that India has conducted too few tests compared to its population size [^30^]. However, a smaller number of testing may not be the only reason behind the low number of COVID-19 confirmed cases in India so far. India has taken many preventive measures to combat COVID-19 in much earlier stages compared to other countries, including nationwide lockdown from 25 March 2020. Apart from the lockdown, people have certain conjectures about possible reasons behind India’s relative success, e.g., measures like the travel ban relatively early, use of Bacille Calmette-Guerin (BCG) vaccination to combat tuberculosis in the population that may have secondary effects against COVID-19 [^31,32^], exposure to malaria and antimalarial drugs [^33^], hot and humid weather slowing the transmission, and so on [^34,35^]. However, as of now, there is no concrete evidence to support these conjectures, although some clinical trials are currently underway to investigate some of these [^36^].

Note that India may have seen fewer COVID-19 cases till now, but the war is not over yet. There are many states like Maharashtra, Delhi, Madhya Pradesh, Rajasthan, Gujarat, Uttar Pradesh, and West Bengal, who are still at high risk. These states may see a huge jump in confirmed COVID-19 cases in the coming days if preventive measures are not implemented properly. On the positive side, Kerala has shown how to effectively “flatten” or even “crush the curve” of COVID-19 cases. We hope India can be free of COVID-19 with a strong determination in policies as already shown by the central and respective state Governments.

There are a few works that are based explicitly on Indian COVID-19 data. Das [^37^] has used the epidemiological model to estimate the basic reproduction number at national and some state levels. Ray et al. [^38^] used a predictive model for case-counts in India. They also discussed hypothetical interventions with various intensities and provided projections over a time horizon. Both the articles have used SIR (susceptible-infected-removed) model (or some extensions) for their analysis and prediction. As we discussed earlier, considering the great diversity in every aspect of India, along with its vast population, it would be a better idea to look at each of the states individually. The study of each of the states individually would help decide further actions to contain the spread of the disease, which can be crucial for the specific states only. In this article, we have mainly focused on the SIS model along with the logistic and the exponential models at each state (restricting to only those states with enough data for prediction). The SIS model takes into account the possibility that an infected individual can return to the susceptible class on recovery because the disease confers no long-standing immunity against reinfection. In South Korea, the health authorities discovered 163 patients who tested positive again after a full recovery [^39,40^]. WHO is aware of these reports of patients who were first tested negative for COVID-19 using PCR (polymerase chain reaction) testing and then after some days tested positive again [^41^].

A report based on one particular model can mislead us. Here, we have considered the exponential, the logistic, and the SIS models along with the daily infection-rate (DIR). We interpret the results jointly from all models rather than individually. We expect the DIR to be zero or negative to conclude that COVID-19 is not spreading in a certain state. Even a small positive DIR (say 0.01) indicates that the virus is still spreading in the community, and can potentially increase the DIR anytime. The states without a decreasing trend in DIR and near exponential growth in active infected cases are Maharashtra, Delhi, Gujarat, Madhya Pradesh, Andhra Pradesh, Uttar Pradesh, and West Bengal. The states with an almost decreasing trend in DIR and non-increasing growth in active infected cases are Tamil Nadu, Rajasthan, Punjab and Bihar. The states with a decreasing trend in DIR and decreasing growth in active infected cases in the last few days are Kerala, Haryana, Jammu and Kashmir, Karnataka, and Telangana. States with non-decreasing DIR need to do much more in terms of the preventive measures immediately to combat the COVID-19 pandemic. On the other hand, the states with decreasing DIR can maintain the same status to see the DIR become zero or negative for consecutive 14 days to be able to declare the end of the pandemic.

## Data Availability

I have used publicly available data for this article. The data source has been mentioned in the article. Here, I put the links again: The three primary sources of the data are the Ministry of Health and Family Welfare, India (https://www.mohfw.gov.in), https://www.covid19india.org/, and Wikipedia (https://en.wikipedia.org/wiki/2020_coronavirus_pandemic_in_India#Statistics).

## Appendix

### Statistical Models

#### Exponential Model

A pandemic can show exponential growth at the initial stage. For example, at the early stage, the 2014-15 Ebola epidemic in West Africa had shown a seemingly exponential spread [^20^]. We can write the exponential model as

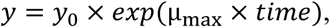

Where *y* is the cumulative confirmed case at a specific time (date), *y_0_* is the initial population, is the maximum growth rate; time is the number of days from first confirmed infection [^42^].

#### Logistic Model

Some pandemics follow an S-shaped curve (sigmoid curve). In other words, the pandemic may start slowly; then, it will increase the growth-rate (infection-rate), and finally, it will flatten the growth-rate over time. The following logistic model can capture that[^42^]

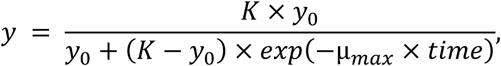

where *K* is the maximum population size; other parameters have the same meaning as in the exponential model.

#### Susceptible Infectious Susceptible (SIS) model

The SIS model is used for a given closed population that is susceptible to a particular disease, is prone to be infected, and communicate the infection within the community [^21^]. It is a time dynamic model with the numbers of susceptible and infected people changing with time according to two different compartments which are characterized by two differential equations:

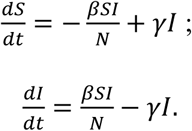

In the above two differential equations, we are trying to observe the rate of change 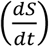 of the susceptible (S) population towards the inflection, and also the rate of change 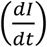 of the infected (I) persons. The model assumes two parameters, namely *β*, which is the average number of contacts per person per unit time, and *γ*, which is obtained as, 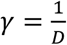, with *D* being the recovery time (specifically, it is the time during which a particular patient can infect others). Here N denotes the total population size with N = S + I.

